# Integrative cross-omics analysis identifies DNA methylation signatures associated with bilateral hippocampal volume, asymmetry and atrophy rate in the general population

**DOI:** 10.1101/2024.12.20.24319418

**Authors:** Dan Liu, Valentina Talevi, Juliana F. Tavares, Ruiqi Wang, Mohammed A Imtiaz, Konstantinos Melas, Alexander Teumer, Katharina Wittfeld, Robert F Hillary, Dina Vojinovic, Marian Beekman, Nicola J Armstrong, Santiago Estrada, Henry Völzke, Robin Bülow, Natalie A. Royle, Joanna M. Wardlaw, Wei Wen, Perminder S. Sachdev, Karen A. Mather, P. Eline Slagboom, Simon R. Cox, Hans Jörgen Grabe, Qiong Yang, N. Ahmad Aziz, Monique M.B. Breteler

**Affiliations:** Population Health Sciences, German Center for Neurodegenerative Diseases (DZNE), Bonn, Germany; Department of Biostatistics, Boston University School of Public Health, Boston, Massachusetts, USA; Department of Psychiatry and Psychotherapy, University Medicine Greifswald, Greifswald, Germany; German Centre for Cardiovascular Research (DZHK), Partner Site Greifswald, Greifswald, Germany; Lothian Birth Cohorts, Department of Psychology, The University of Edinburgh, UK; Molecular Epidemiology, Department of Biomedical Data Science, Leiden University Medical Center, Leiden, The Netherlands; Mathematics and Statistics, Curtin University, Perth, Australia; Artificial Intelligence in Medical Imaging, German Center for Neurodegenerative diseases (DZNE), Bonn, Germany; Institute for Community Medicine, University Medicine Greifswald, Greifswald, Germany; Institute of Diagnostic Radiology and Neuroradiology, University Medicine Greifswald, Greifswald, Germany; Brain Research Imaging Centre, The University of Edinburgh, Edinburgh, UK; Center for Healthy Brain Ageing, Discipline of Psychiatry and Mental Health, School of Clinical Medicine, University of New South Wales, Sydney, Australia; Max Planck Institute for Biology of Aging, Cologne, Germany; German Center for Neurodegenerative Diseases (DZNE), Partner Site Rostock/Greifswald, Greifswald, Germany; The Framingham Heart Study, Framingham, Massachusetts, USA; Department of Neurology, Faculty of Medicine, University of Bonn, Bonn, Germany; Institute for Medical Biometry, Informatics and Epidemiology (IMBIE), Faculty of Medicine, University of Bonn, Bonn, Germany

## Abstract

The hippocampus exhibits volumetric differences between the left and right hemispheres (LHCV and RHCV) and asymmetry, yet the molecular mechanisms underlying these features remain unclear. In this study, we performed a meta-analysis of epigenome-wide association studies (EWAS) across six population-based cohorts (n=8,156; 53% women; mean age: 60.7 years) to identify blood-based methylation signatures associated with LHCV, RHCV, and hippocampal asymmetry. Integrative cross-omics analyses using individual-level genetic, methylation, and gene expression data (n > 2,624 participants from the Rhineland Study) revealed 15 CpG/DMR-gene expression pairs linked to LHCV and 18 pairs to RHCV, implicating pathways involved in neuronal differentiation and immune processes. Notably, baseline methylation at these loci predicted bilateral hippocampal atrophy rates, explaining over 10% of the variation. Four CpGs were consistently associated with hippocampal asymmetry cross-sectionally and longitudinally, exhibiting sex-specific differences. Additionally, adherence to healthy dietary patterns was associated with these methylation signatures, highlighting modifiable influences on hippocampal health and atrophy.

## Introduction

The human hippocampus, a bilateral brain structure within the middle temporal lobe, is essential for episodic memory, spatial navigation, as well as other cognitive functions.^1^ Large-scale neuroimaging analyses have revealed that the hippocampus is one of the most consistently and robustly altered gray matter structures involved in several neurodegenerative and neuropsychiatric disorders, including Alzheimer’s disease,^2^ major depressive disorder,^3^ schizophrenia,^4,5^ and attention deficit hyperactivity disorder.^6^ Previous studies have revealed both structural and functional hippocampal asymmetries.^7–14^ Specifically, in humans, left hippocampal volume (LHCV) is slightly smaller than right hippocampal volume (RHCV), and task-related activity may be localized to only one hemisphere.^12,15,16^ This lateralization may enable the left and right hippocampus to support complementary functions in human episodic memory and navigation. For instance, the left hippocampus tends to dominate when semantic information is most relevant to the task, while the right hippocampus takes precedence when spatial information is more important.^7,10^ Additionally, prior studies have suggested that the left and right hippocampus are affected differently by the aging process,^17^ and pathological factors may induce further asymmetry.^15,16^ Nonetheless, the molecular characteristics underlying the (age-related) reduction of LHCV and RHCV and increase in hippocampal asymmetry remain largely unknown in the general population.

Genome-wide association studies (GWAS) have discovered a number of genetic variants related to LHCV, RHCV, and hippocampal asymmetry.^12,13,18^ However, the majority of the identified genetic loci are located in non-coding genomic regions, suggesting they influence transcriptional regulation rather than protein-coding sequences. This transcriptional regulation may be significantly affected by DNA methylation changes,^19^ which involve the addition of a methyl group (CH_3_) to the genomic DNA and is responsive to both internal and external stimuli, such as environmental changes and aging.^20^ Recent studies have demonstrated that CpG methylation-dependent transcriptional regulation is a widespread phenomenon, which could provide molecular links between complex traits.^20^ In addition, DNA methylation levels may influence the binding of transcription factors to DNA, which is essential for tuning of gene expression levels.^21,22^ Thus, the interplay between genetics, DNA methylation, and gene expression may contribute to the variability in LHCV, RHCV, and hippocampal asymmetry across the adult lifespan.

Indeed, a previous study of 649 Alzheimer’s Disease Neuroimaging Initiative participants and mouse models using CRISPR-Cas9 (epi)genome-editing techniques revealed a causal relationship between the presence of a single nucleotide polymorphism (SNP) (rs1053218) and hypermethylation of a specific CpG site (cg26741686), leading to higher *ANKRD37* gene expression and reduction of mean hippocampal volume.^23^ An epigenome-wide meta-analysis including 3,337 samples found two CpGs and three DNA methylated regions, which influenced gene expression and were associated with mean hippocampal volume.^11,24^ However, previous investigations primarily focused on total or mean hippocampal volume from heterogeneous and relatively small samples (N< 3,400), which also included patients with neurological and neuropsychological disorders. Therefore, the DNA methylation signatures of LHCV, RHCV, and hippocampal asymmetry in the general population largely remain unknown. Moreover, previous studies utilized the HumanMethylation450 array (HM450K), which measures 450,000 CpGs, or assessed specific CpGs, limiting their capability to detect other potentially relevant CpGs. In contrast, the HumanMethylationEPIC array (HM850K) encompasses a substantially wider unparalleled coverage of 850,000 CpGs across the genome,^25^ which could help to identify novel methylation markers related to LHCV, RHCV, and hippocampal asymmetry separately. Importantly, few studies have comprehensively assessed the functional characterization of these methylation differences at the genetic and transcriptional levels, potentially missing key regulatory mechanisms that may not be apparent when studying each molecular layer in isolation. Additionally, it remains unexplored whether these cross-sectional changes are associated with hippocampal atrophy and if modifiable lifestyle factors have an impact on them.

In this study, we meta-analyzed epigenome-wide association analyses (EWAS) from six population-based studies comprising 8,156 samples to uncover the methylation signatures of LHCV, RHCV, and hippocampal asymmetry separately. To evaluate whether these associations were specific to the hippocampus or due to global gray matter changes, we also performed EWASs of left hemisphere gray matter volume (LGMV), right hemisphere gray matter volume (RGMV), and global gray matter asymmetry. By leveraging concomitant individual-level genetics, DNA methylation, and gene expression data from participants of the Rhineland Study, we performed integrative cross-omics analyses. These analyses aimed to delineate the complex regulatory connectivity between genetics, methylation, gene expression, and transcription factor binding sites, prioritizing potential molecular mechanisms contributing to structural and functional differences in the hippocampus. To facilitate understanding of the clinical implications of the identified methylation signatures, we further assessed their association various dietary patterns and with longitudinal changes in LHCV, RHCV and hippocampal asymmetry.

## Results

### Study sample characteristics

Our workflow to identify novel methylation signatures of LHCV, RHCV, and hippocampal asymmetry is presented in **Fig. 1 & Fig. S1**. The sample characteristics are presented in **Table 1**. Our meta-analysis included 8,156 samples from six population-based cohort studies. Details about the included studies are provided in **Table S1-S3.** The mean age of the participants of the Rhineland Study, SHIP-Trend, FHS and LLS ranged from 50.4 years to 58.5 years, while participants from LBC1936 and OATS were older (LBC1936 mean age: 72.6 years, OATS mean age: 70.5 years). Sex ratios were balanced in all studies. On average, LHCV was slightly smaller than RHCV in all cohorts (**Table 1**), and both decreased with increasing age after adjusting for estimated total intracranial volume (eTIV) (**Fig. S2**). The mean value of hippocampal asymmetry ranged from −0.0058 in FHS to −0.0400 in LBC1936, and it slightly increased with age (**Fig. S2**).

**Fig. 1.**
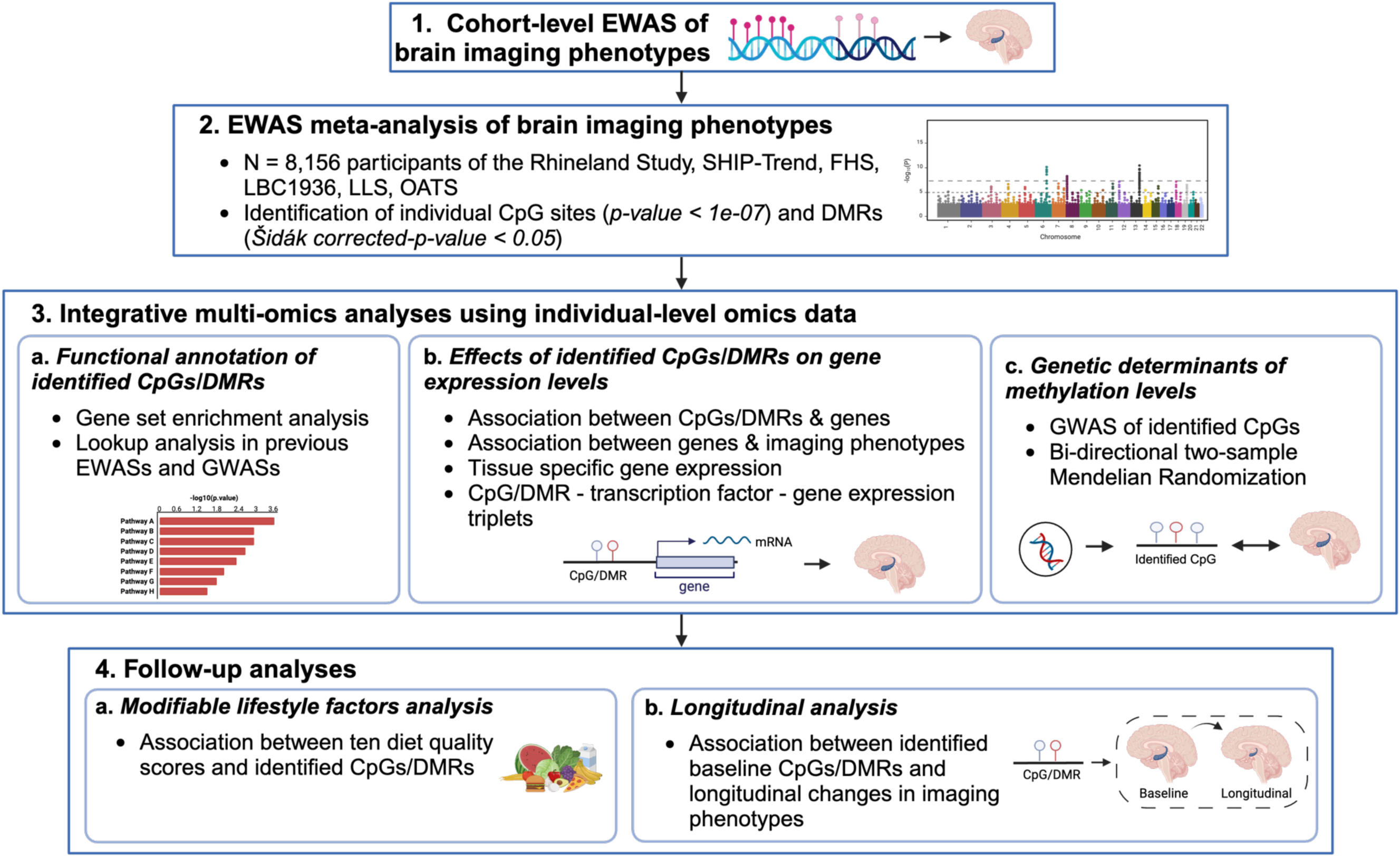
Workflow for identifying DNA methylation signatures associated with bilateral hippocampal volume and asymmetry. The integrative multi-omics analyses and follow-up were performed using individual level genetics, DNA methylation, gene expression, dietary and longitudinal brain imaging data measured in the same participants from the Rhineland Study. Abbreviations: EWAS, epigenome-wide association study; DMR, differentially methylated region; GWAS, genome-wide association study.

**Table 1:** Characteristics of the participating cohorts.

| **Cohort** | **Rhineland Study** | **SHIP-Trend** | **FHS** | **LBC1936** | **LLS\*** | **OATS** |
| --- | --- | --- | --- | --- | --- | --- |
| Sample size, N | 4568 | 451 | 2317 | 560 | 173 | 87 |
| Age (years), mean (SD) | 54.4<br>(13.4) | 50.4<br>(13.4) | 57.7<br>(13.1) | 72.6<br>(0.7) | 58.5<br>(6.6) | 70.5<br>(5.4) |
| Age range (years) | 30 - 92 | 22 - 79 | 25 - 90 | 71 - 74 | 40 -79 | 65 - 83 |
| Women, % | 57.4 | 55.5 | 47.1 | 46.6 | 56.7 | 58.6 |
| Estimated total intracranial volume (cm<sup>3</sup>), mean (SD) | 1550.00<br>(148.00) | 1573.29<br>(160.01) | 1289.19<br>(132.60) | 1451.20<br>(143.54) | 1426.61<br>(125.43) | 1442.510<br>(171.671) |
| Left hippocampal volume (cm<sup>3</sup>), mean (SD) | 3.84<br>(0.44) | 3.88<br>(0.42) | 3.36<br>(0.39) | 3.07<br>(0.44) | 5.17<br>(0.59) | 3.66<br>(0.42) |
| Right hippocampal volume (cm<sup>3</sup>), mean (SD) | 4.00<br>(0.48) | 4.04<br>(0.43) | 3.40<br>(0.38) | 3.33<br>(0.44) | 5.29<br>(0.53) | 3.75<br>(0.44) |
| Hippocampal asymmetry index, mean (SD) | -0.0195<br>(0.0276) | -0.0200<br>(0.0254) | -0.0058<br>(0.0290) | -0.0400<br>(0.0400) | -0.0112<br>(0.0497) | -0.0118<br>(0.0304) |
| Left hemispheric gray matter volume (cm<sup>3</sup>), mean (SD) | 256.00<br>(26.60) | 257.51<br>(26.91) | 267.65<br>(28.18) | 201.54<br>(18.77) | - | 193.62<br>(19.13) |
| Right hemispheric gray matter volume (cm<sup>3</sup>), mean (SD) | 257.00<br>(26.20) | 258.51<br>(27.08) | 267.37<br>(27.88) | 202.95<br>(19.04) | - | 195.15<br>(18.98) |
| Hemispheric gray matter asymmetry index, mean (SD) | -0.00136<br>(0.00536) | -0.00196<br>(0.00458) | 0.00047<br>(0.0077) | -0.0030<br>(0.0100) | - | -0.0040<br>(0.0072) |
| Methylation array type | HM850K | HM850K | HM450K | HM450K | HM450K | HM450K |

Abbreviation: SHIP-Trend, the Study of Health in Pomerania; FHS, the Framingham Heart Study; LBC1936, the Lothian Birth Cohort 1936; LLS, the Leiden Longevity Study; OATS, the Older Australian Twin Study; HM850K, Illumina Infinium MethylationEPIC BeadChip array; HM450K, Illumina Infinium Methylation450K BeadChip array; SD, standard deviation.
\*Gray matter volumes are not available in LLS.

### Specific methylation signatures were associated with hippocampal volume and asymmetry at individual CpG and differentially methylated region level

The meta-analysis revealed that five CpGs were associated with LHCV, nine CpGs were associated with RHCV, and one CpG was associated with hippocampal asymmetry (**Fig. 2A**). The top CpGs remained the same after further adjustment for handedness (**Fig. S3**). In addition, we found sex differences in single CpG associations with hippocampal asymmetry (p-values for z-test < 0.05) (**Fig. 2B**). Intriguingly, minimal overlap was observed in the associated CpGs and their mapped genes among LHCV, RHCV, and hippocampal asymmetry. Moreover, there was no overlap between CpGs associated with hippocampal traits and global gray matter traits (**Fig. S4**), indicating that the effects were specific to the hippocampus rather than due to global gray matter changes.

**Fig. 2.**
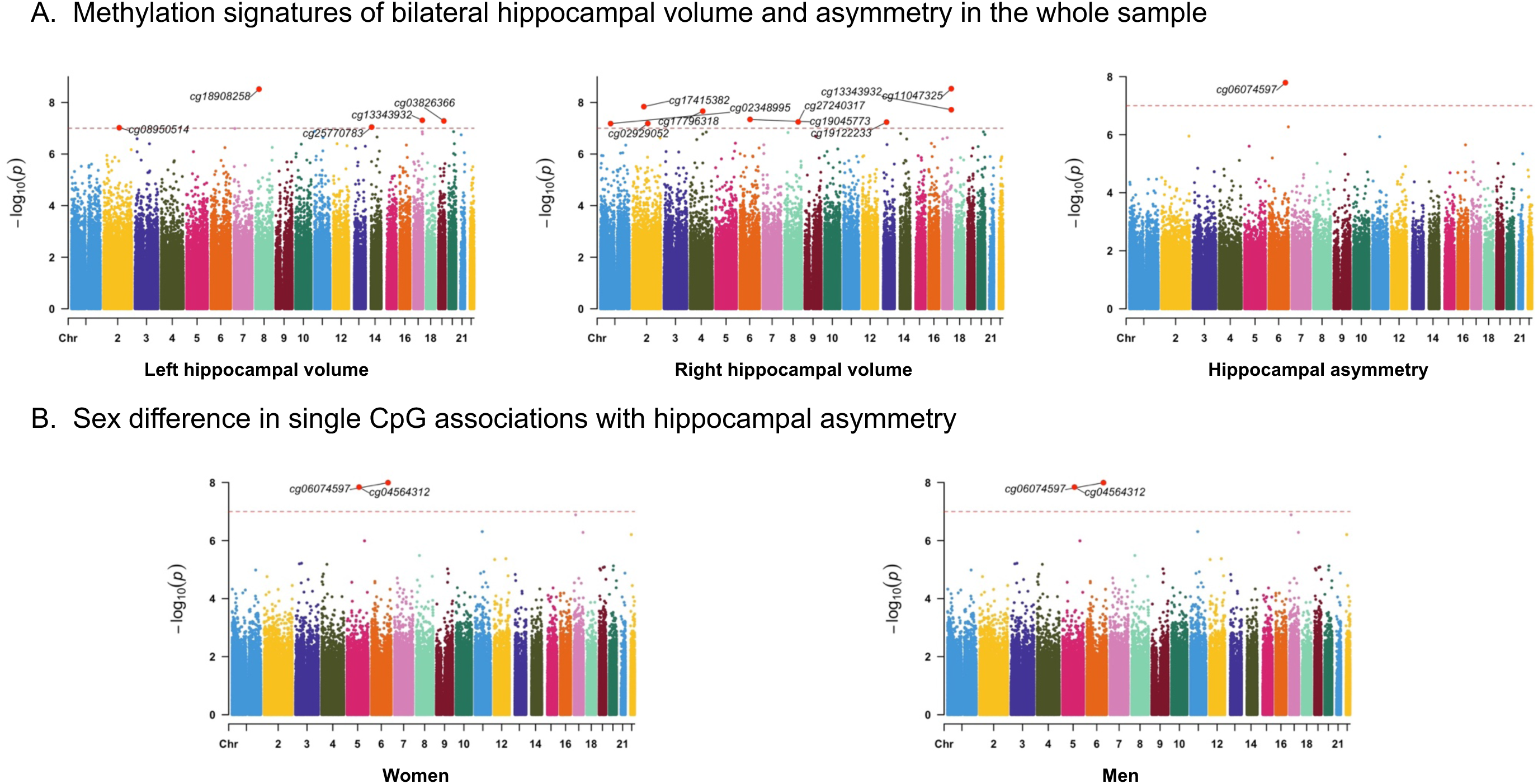
Manhattan plots of the epigenome-wide meta-analyses of bilateral hippocampal volume and asymmetry. Results were plotted as negative log-transformed p-values (y-axis) across the genome (x-axis). The red horizontal line represents the epigenome-wide significance at 1.0 x10^-7^. Linear models were adjusted for age, sex (not for panel B), batch effects, blood cell proportions, first ten genetic principal components (to account for population stratification), smoking status, and education. Models for bilateral hippocampal volumes were additionally adjusted for estimated total intracranial volume. All the epigenome-wide significant CpGs were annotated.

Accounting for the joint effect of spatially correlated CpGs clustered in specific genomic regions, we identified 262, 246, and 16 differentially methylated regions (DMRs) associated with LHCV, RHCV, and hippocampal asymmetry, respectively (all Šidák corrected-p-value < 0.05, number of consecutive probes ≥ 2). Importantly, CpGs identified from the individual CpG analysis were also detected in the DMR analysis (**Table S4**). Additionally, 31.4% of DMRs overlapped between LHCV and RHCV, less than 10% of DMRs overlapped between hippocampal traits and global gray matter traits, and only two DMRs associated with hippocampal asymmetry were also related to LHCV and/or RHCV (**Fig. S5**).

### *In silico* replication

Previous EWAS analyses identified three CpGs (cg17858098, cg26927218, and cg26741686) and two DMRs associated with mean hippocampal volume.^24^ In our study, these two DMRs were also associated with LHCV/RHCV (**Table S5**). Higher levels of cg17858098 methylation were previously associated with larger mean hippocampal volume. However, we found that higher levels of cg17858098 methylation tended to be associated with larger RHCV, but smaller LHCV (**Table S5**). Although cg26927218 was not present in our study, the previous EWAS found it to be associated with *BAIAP2* gene expression. Our study showed that higher methylation levels of the RHCV-related CpGs, cg13343932 and cg110477325, were associated with higher *BAIAP2* gene expression and smaller RHCV.

### Gene set enrichment analysis for the identified methylation signatures

These identified methylation signatures largely converged on pathways including, but not limited to, nervous system development, neuron differentiation, generation of neurons, and neuronal morphogenesis (**Fig. S6** & **S8**). Moreover, these distinct CpGs/mapped genes were involved in trait-specific biological pathways (**Fig. S7**). For instance, cell function-related pathways were associated with LHCV-related CpGs, neuron projection regulation and post-synapse organization were associated with RHCV-related CpGs, whereas localization-related pathways were associated with hippocampal asymmetry (**Fig. S7**).

Additionally, LHCV- and RHCV-related DMR-mapped genes have been linked to hippocampal volume (i.e., total and hippocampal tail volume) in previous GWAS analyses, confirming their (epi)genetic function in hippocampus. These DMR-mapped genes have also been linked to a wide range of neurodegenerative and psychiatric disorders, as well as to CSF Aβ_1-42_ levels, t-tau/ Aβ_1-42_ ratio, and p-tau181p levels (**Table S6**). Meanwhile, it is worth noting that LHCV-related DMR-mapped genes have been previously linked to global cognition, processing speed, and word reading, where semantic information is more relevant, whereas RHCV-related DMR-mapped genes have been previously linked to logical memory and reaction time (**Table S6**).

### Integrative multi-omics analyses reveal functional implications of methylation differences in hippocampal volume and asymmetry

To better understand the functional roles of the identified CpGs and DMRs, we performed several integrative omics analyses using indivdual-level DNA methylation, gene expression, and genotype data from the same participants in the Rhineland Study. The overview of the datasets used in the analyses is presented in **Fig. S1**.

#### Associations of methylation levels of significant CpGs and DMRs with expression of nearby genes and corresponding phenotypes

At the individual CpG level, 14 top CpGs were mapped to 13 genes. Nine out of these 13 mapped genes were present in our gene expression data, and we detected five significant CpG-gene expression pairs. Among the 1,543 *cis*-genes for the top CpGs, 620 genes were present in our data. All five LHCV-related CpGs showed associations with their *cis*-gene expressions, resulting in 81 significant CpG-gene expression pairs. Notably, the expression levels of six genes (i.e. *LGALS3BP, CD300LB, USP36, TRAJ19, GEMIN7,* and *NUGGC*) were also associated with LHCV (**Fig. 3A**). Similarly, all nine RHCV-related CpGs showed associations with the expression levels of their *cis*-genes, yielding 100 significant CpG-gene expression pairs. Among them, the expression levels of nine genes (i.e. *LGALS3BP, CD300LB, MCOLN2, TGFBR3, SNORD89, BAIAP2, SLC16A5, SYNGR2,* and *CD300C* gene) were also associated with RHCV (**Fig. 3A**). Interestingly, only two *cis*-genes of cg13343932, *LGALS3BP* and *CD300LB*, overlapped between LHCV and RHCV, indicating that different molecular pathways (CpG-gene expression-LHCV/RHCV) affect LHCV and RHCV. Additionally, hippocampal asymmetry-related cg06074597 was associated with the expression of three genes; however, only *SASH1* gene expression was borderline significantly associated with hippocampal asymmetry (beta: 0.042, 95%CI: −0.001 – 0.086).

**Fig. 3.**
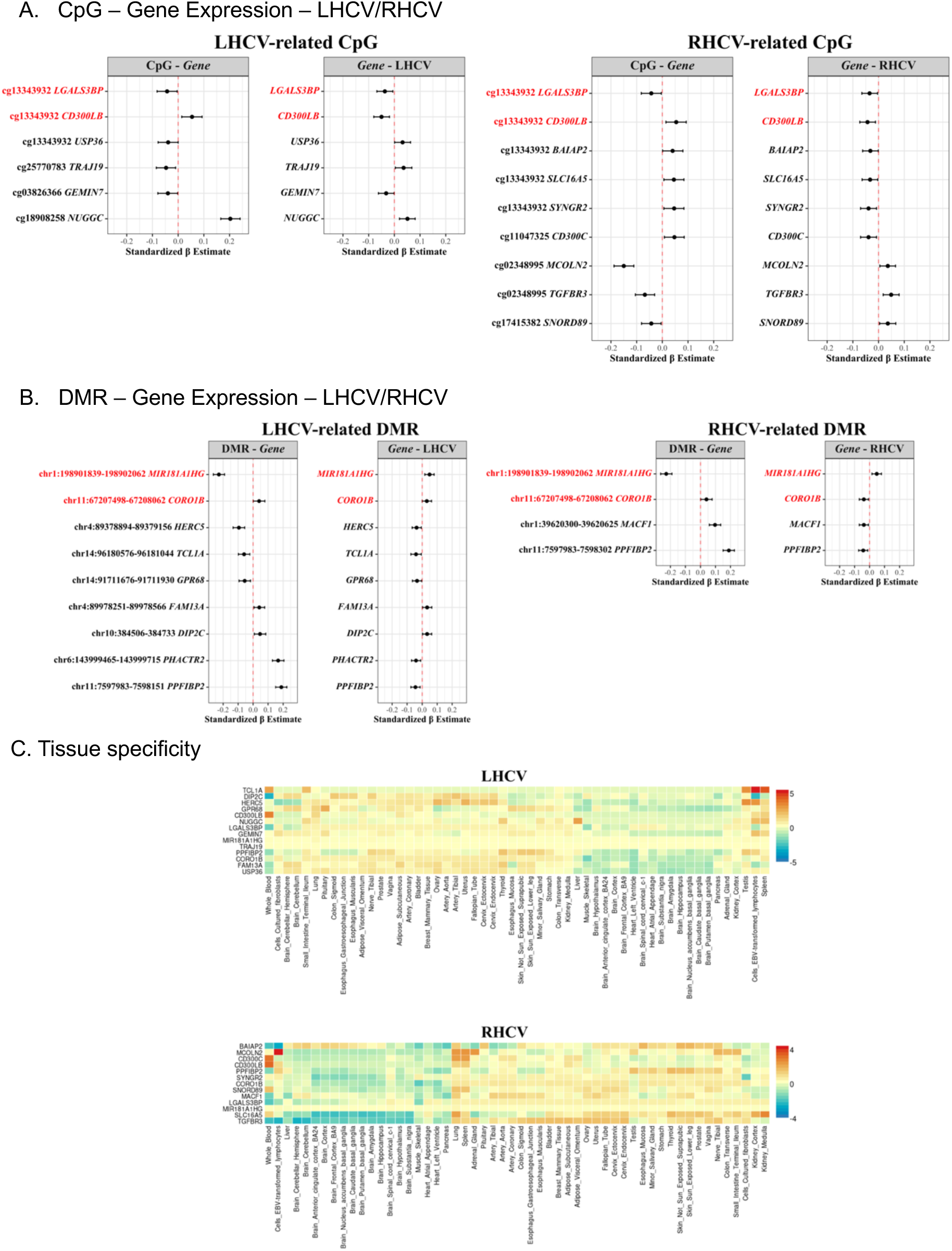
The associations between identified methylation signatures, gene expression, and bilateral hippocampal volume. (A) Forest plots showing the associations between identified methylation signatures and the mapped or *cis*-genes and the associations between these candidate genes and LHCV/RHCV. The dot represents the mean effect and the horizontal line shows the 95% CI. Genes overlapping between LHCV and RHCV are shown in red. (B) Forest plots showing the associations between identified DMR signatures and the mapped genes and the associations between these candidate genes and LHCV/RHCV. The dot represents the mean effect and the horizontal line shows the 95% CI. Genes overlapping between LHCV and RHCV are shown in red. (C) Heatmaps showing the expression of candidate genes across different tissues. The heatmaps were generated using FUMA v1.6.1 and gene expression data from GTEx v8 54 tissue types. Abbreviations: CI, confidence interval; LHCV and RHCV, left and right hippocampal volumes; DMR, differentially methylated region.

Out of 389 DMR-mapped genes, 183 were present in our data. The DMRs encompassing the top CpGs showed slightly stronger associations with gene expression compared to individual CpGs. We found that 55 of the 134 LHCV-related DMRs were associated with their mapped gene expression. Among these, expression levels of nine genes were also associated with LHCV (**Fig. 3B**). Similarly, 46 of the 115 RHCV-related DMRs were associated with their mapped gene expressions, with four of these genes associated with RHCV (**Fig. 3B**). Additionally, four out of five hippocampal asymmetry-related DMRs were associated with the expression of their mapped genes; however, none of these genes were associated with hippocampal asymmetry. Interestingly, only *MIR181A1HG* and *CORO1B* overlapped between LHCV and RHCV, while no overlaps were observed with hippocampal asymmetry (**Fig. 3B**).

Furthermore, we identified sex-specific differences in CpG–gene expression associations related to hippocampal asymmetry. In women, cg16747427 and ch.6.169008488F were associated with hippocampal asymmetry (**Fig. 2B)**. cg16747427 was associated with 11 *cis*-gene expression levels, among which the expression *of ABT1* gene was associated with hippocampal asymmetry in women (beta: −0.058, 95%CI: −0.116 – 0.0003). In men, cg04564312 and cg06074597 were associated with hippocampal asymmetry (**Fig. 2B)**, and cg04564312 was associated with the *RGMB* gene expression (beta: −0.064, 95%CI: −0.12 – 0.008).

We further investigated whether gene expression mediates the relationship between CpGs/DMRs and their associated traits. Our analysis revealed significant mediation of the associations between the DMRs chr11.7597983.7598302 and chr1.39620300.39620625 with RHCV through the gene expression of *PPF1BP2* (25.7% mediation) and *MACF1* (16.9% mediation) (**Table S7**). The mediation effects for other CpG/DMR-gene-trait pairs were not statistically significant, likely due to the relatively smaller sample size (n = 1,800 participants with available methylation, gene expression, and brain MRI data).

#### Tissue specificity

To assess the brain-specfic relevance of these findings, we compared the expression levels of the identified genes across different tissues, especially between blood and hippocampus (**Fig. 3C**). The majority of LHCV-related genes exhibited similar expression levels in blood and hippocampus. Likewise, most RHCV-related genes showed comparable expression levels between these two tissues.

#### Hippocampus-related CpGs/DMRs are associated with putative transcription factors and target genes

We found that the majority of the identified methylation signatures were located in the promoter regions and the gene body (**Fig. S9**). Importantly, almost all of them are gene regulatory elements, including DNase hypersensitive sites (DHSs), transcription factor binding sites (TFBS), or showed evidence for open chromatin (**Table S8**), supporting their dynamic interaction with transcription factors (TFs) in gene expression regulation.

To further assess whether these methylation signatures influence the regulatory activities of transcription factors on target gene, we next performed an integrative analysis of DNA methylation, TF binding, and gene expression data to uncover CpG/DMR-TF-target gene triplets. At the CpG level, we discovered the cg25770783-FOXO4 transcription factor-*DHRS1* gene expression triplet, in which LHCV-related DNA methylation at cg25770783 appeared to attenuate the activation of the target gene *DHRS1* by the transcription factor FOXO4. Specifically, in participants with low cg25770783 methylation levels, higher FOXO4 activity corresponded to higher activation of the *DHRS1* gene (p-value = 8.13 × 10^−7^). Conversely, when cg25770783 methylation was high, the target gene was relatively independent of transcription factor activities (**Fig. S10A**).

At the DMR level, several additional TFs interact with LHCV-, RHCV-, and hippocampal asymmetry-related DMR methylation to jointly regulate target gene expression. Notably, ZNF354C was affected by several DMRs across traits in regulating the gene expression levels of multiple genes. Among them, methylation at DMR chr1:174843523-174843971 and transcription factor ZNF354C jointly regulate *GPR52* gene expression related to LHCV, and methylation at DMR chr15:31685635-31685823 and transcription factor ZNF354C jointly regulate *OTUD7A* gene expression related to RHCV (**Fig. S10B**). Both DMRs are within gene regulatory elements (**Table S8**).

#### Genetic contribution to the methylation status of the identified CpGs and bidirectional two sample Mendelian Randomization (MR) associations

Our GWAS analyses identified 595, 228, and 154 genome-wide significant SNPs associated with three LHCV-related CpGs (**Fig. S11A**). For the nine RHCV-related CpGs, we identified 3,188 genome-wide significant SNP-CpG pairs (**Fig. S11B**). After linkage disequilibrium (LD)-clumping, the independent SNPs for each CpG were used as genetic instrumental variables in the subsequent MR analyses.

The bidirectional two-sample MR revealed a significant causal relationship between higher methylation at cg19045773 and smaller RHCV (inverse variance-weighted (IVW)-based beta = −0.945, standard error (SE) = 0.417, p-value = 0.023, **Fig. S12A**). The effect size and direction were consistent across MR models, with no evidence of heterogeneity (Cochran’s Q test, p-value = 0.701) or horizontal pleiotropy (MR Egger regression intercept p-value = 0.399). Additionally, methylation at cg19045773 was associated with expression levels of the *cis*-genes *RRAGD* and *ANKRD6 (***Fig. S12C)**.

We also found evidence suggesting that higher methylation at cg02929052 was causally associated with larger RHCV (IVW beta = 2.063, SE = 0.956, p-value = 0.03, **Fig. S12A**). There was no evidence for heterogeneity (Cochran’s Q test, p-value = 0.331). However, since only two genetic instrumental variables were available, MR analysis with alternative methods could not be performed. In addition, higher cg02929052 methylation was associated with lower *AMMECR1L* and *UGGT1* gene expression (**Fig. S12C**).

#### The association between diet quality scores and identified methylation signatures

Leveraging individual-level dietary intake data from the Rhineland Study, we observed that higher adherence to healthy dietary patterns—such as Plant-based Diet (PDI), Alternate Healthy Eating Index (AHEI), Nordic, Dietary Approaches to Stop Hypertension (DASH), and EAT-Lancet diets—was consistently associated with higher methylation levels at several CpGs, specifically cg133343932 and cg11047325. Conversely, adherence to unhealthy dietary patterns (e.g., Dietary Inflammatory Index (DII) and unhealthful PDI) was linked to lower methylation levels at the same CpGs (**Fig. 4A**).

**Fig. 4.**
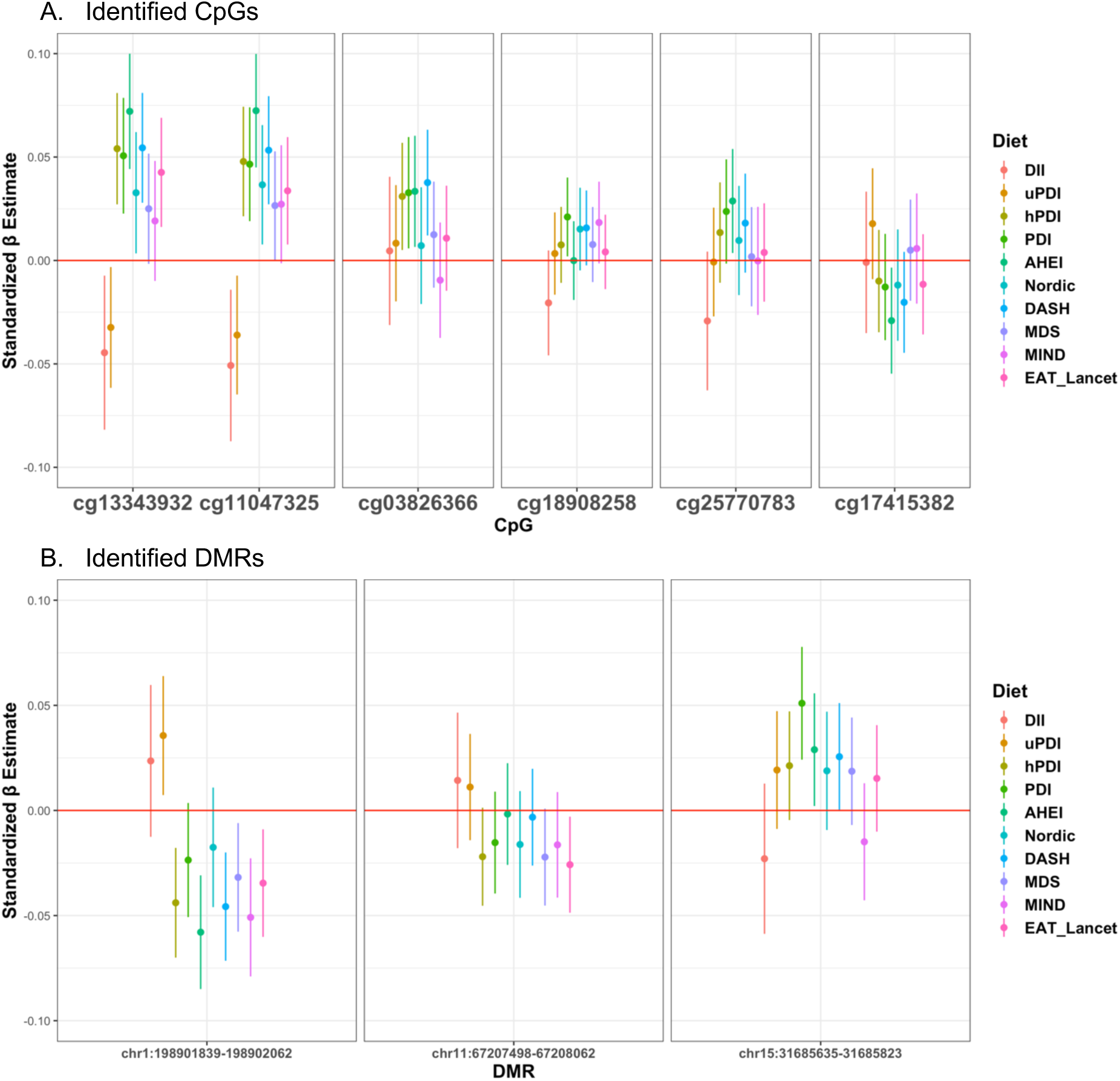
The association between dietary patterns and identified methylation signatures. (A) Forest plots showing the associations between dietary patterns and the identified individual CpGs. The dot represents the mean effect and the vertical line shows the 95% CI. Color indicates different dietary patterns. (B) Forest plots showing the associations between dietary patterns and the identified DMR signatures. The dot represents the mean effect and the vertical line shows the 95% CI. Color indicates different dietary patterns. Abbreviations: MDS, Mediterranean Diet Score; DASH, Dietary Approaches to Stop Hypertension; MIND, Mediterranean-DASH Intervention for Neurodegenerative Delay; AHEI, Alternate Healthy Eating Index score; PDI, Plant-based Diet Index; hPDI, Healthful Plant-based Diet Index; uPDI, Unhealthful Plant-based Diet Index; DII, Dietary Inflammatory Index.

Similarly, we found that greater adherence to multiple dietary patterns was associated with lower methylation levels at LHCV/RHCV common-related DMRs (**Fig. 4B**), particularly those mapped to *MIR181A1HG* or *CORO1B*. Moreover, PDI and AHEI diets were associated with a RHCV-related DMR mapped to the *KLF13* gene (**Fig. 4B**).

#### Association of identified baseline methylation signatures with longitudinal changes in imaging measures

Among the CpGs cross-sectionally associated with LHCV, higher baseline methylation at cg13343932 and cg25770783 was significantly associated with slower left hippocampal volume loss over time, while higher baseline methylation at cg18908258 was associated with faster left hippocampal volume loss over time (**Fig. 5A**). For instance, the average annual LHCV change rate was −15.83 mm^3^ in the linear mixed model for cg25770783. However, when baseline cg25770783 methylation was one standard deviation higher, the estimated average annual change rate improved to −14.74 mm^3^, reflecting a 7.5% reduction (1.1 mm^3^) in yearly left hippocampal volume loss. Notably, these three CpGs together explained 15.6% of the variation in left hippocampal volume loss rates. Similarly, among the six CpGs cross-sectionally associated with RHCV, higher baseline methylation at cg11047325, cg13343932 and cg02348995 was significantly associated with slower right hippocampal volume loss over time, whereas higher baseline methylation at cg02929052 was associated with faster right hippocampal volume loss over time (**Fig. 5A**). These four CpGs together explained 10.9% of the variation in right hippocampal volume loss rates.

**Fig. 5.**
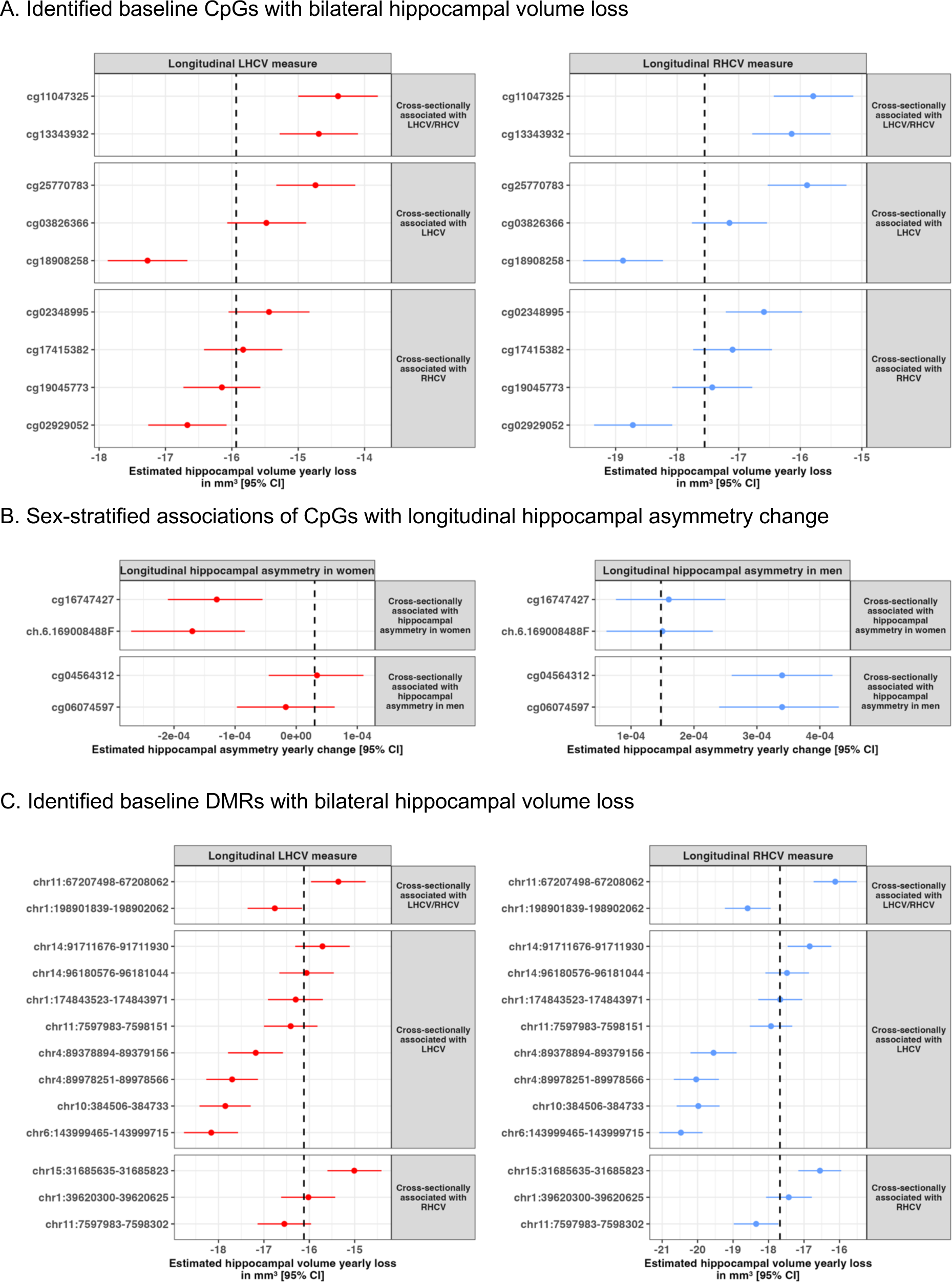
Association between identified baseline methylation signatures and longitudinal change in imaging measures. The dot represents the mean annual change rate in imaging measures per standard deviation increase in baseline methylation, while the vertical line indicates the 95% confidence interval. The dashed line in each plot indicates the yearly change rate of each measure when CpG expression was equal to the population mean, averaged across all CpGs included in the plot. To facilitate plotting, different axis scales have been used for hippocampal volume change, and asymmetry change. Abbreviations: LHCV and RHCV, left and right hippocampal volumes; DMR, differentially methylated region; CI, confidence interval.

Furthermore, in women, higher baseline methylation at cg16747427 and ch.6.169008488F, which were cross-sectionally associated with less hippocampal asymmetry, was significantly associated with decreased hippocampal asymmetry over time. These two CpGs together explained 4.4% of the variation in hippocampal asymmetry change rates in women. In men, higher baseline methylation at cg04564312 and cg06074597, which were cross-sectionally associated with more hippocampal asymmetry, was associated with increased hippocampal asymmetry over time (**Fig. 5B**). These two CpGs together explained 30% of the variation in hippocampal asymmetry change rates in men.

At the DMR level, among the DMRs cross-sectionally associated with LHCV, higher baseline methylation at five DMRs was significantly associated with slower left hippocampal volume loss over time, and higher baseline methylation at one DMR was associated with faster left hippocampal volume loss over time. For RHCV, higher baseline methylation at two DMRs was associated with slower right hippocampal volume loss over time, and higher baseline methylation at another two DMRs was significantly associated with faster right hippocampal volume loss over time (**Fig. 5C**).

## Discussion

By leveraging epigenome-wide association studies across several population-based cohorts, we identified blood-based methylation signatures associated with LHCV, RHCV and hippocampal asymmetry throughout the adult lifespan. These signatures mainly converged on pathways involved in neuronal differentiation and development. Integrative cross-omics analyses using deep-phenotyping data from the Rhineland Study revealed 15 and 18 CpG/DMR–gene expression pairs associated with LHCV and RHCV, respectively, with four CpGs associated with hippocampal asymmetry displaying notable sex-specific differences. Importantly, various dietary patterns were associated with methylation levels at these loci, and baseline methylation signatures at these loci predicted longitudinal changes in hippocampal volume and asymmetry, collectively explaining >10% of the variation in bilateral hippocampal atrophy change rates.

Our functional omics analyses revealed potential molecular mechanisms through which gene methylation influences bilateral hippocampal atrophy. A notable group of implicated genes was immune-related, while another group was involved in neuronal structure and synaptic function. For example, methylation changes at cg133443932 and cg11047325 and DMRs (chr1:198901839-198902062 and chr11:67207498-67208062) and their associated cis-genes - *LGALS3BP, CD300LB, MIR181A1HG, and CORO1B -* were significantly associated with both left and right hippocampal volume. Intriguingly, baseline methylation at these loci was predictive of bilateral hippocampal atrophy rates, explaining a considerable proportion of the variation in the age-associated rates of atrophy. Moreover, adherence to healthy dietary patterns was strongly associated with these same methylation signatures, suggesting that dietary intake could modulate the rate of hippocampal atrophy through its effects on DNA methylation.

LGALS3BP (galectin-3 binding protein) is a secreted protein that interacts with various members of the extracellular matrix and is highly enriched in human neural progenitors.^26^ It has been implicated in modulating the immune response, particularly in processes involving natural killer and lymphokine-activated killer cell cytotoxicity.^27^ Notably, extracellular LGALS3BP has been shown to regulate neural progenitor positioning, which may contribute to the development of human cortical complexity.^28^ The CD300 family of molecules regulates a diverse array of immune cell processes through its paired activating and inhibitory receptors. Among these, CD300LB (also known as TREM5 or CLM7) is an activating receptor of the immunoglobulin (Ig) superfamily, predominantly expressed on myeloid cells. CD300LB plays a critical role in modulating immune responses, frequently by interacting with lipid ligands and other cellular components.^26^ Nutritional patterns associated with reduced inflammation and enhanced immune resilience, such as the EAT-Lancet, AHEI, and Nordic diets, may influence the regulation of CD300LB. These diets might enhance the receptor’s capacity to maintain a balanced immune response, contributing to the protection of neural tissue from excessive inflammation. Conversely, diets high in processed foods and pro-inflammatory components could dysregulate CD300LB activity, potentially exacerbating immune-mediated damage in the central nervous system.^29^

The *MIR181A1HG* gene encodes microRNA-181a, which regulates key pathways involved in neurogenesis, synaptic plasticity, and apoptosis.^30^ In addition, miR-181a has been linked to cognitive function and has been proposed as a potential target against cognitive decline.^31^ Dietary patterns have been shown to influence miR-181a expression,^32^ particularly those that are mainly plant-based, which promote the intake of foods rich in antioxidants and anti-inflammatory ingredients (i.e., fruits, vegetables, nuts, etc.) such as the Mediterranean and EAT-Lancet diets. Higher methylation levels in response to these diets could potentially downregulate inflammatory pathways in the hippocampus, protecting against cognitive decline. On the other hand, pro-inflammatory diets, such as those high in processed foods (e.g., high DII scores), may reduce *MIR181A1HG* methylation, possibly amplifying inflammatory signals that can impair hippocampal function and brain plasticity. The *CORO1B* (Coronin 1B) gene is essential for regulating the actin cytoskeleton,^33^ which is crucial for neuronal structure and synaptic function. Proper actin dynamics influence hippocampal architecture and synaptic plasticity—key aspects of learning and memory.

Our findings reveal sex-specific epigenetic associations with hippocampal asymmetry, and point towards potential underlying molecular mechanisms. Specifically, in women, two CpG sites, cg16747427 and ch.6.169008488F, were consistently associated with hippocampal asymmetry both cross-sectionally and longitudinally. Notably, cg16747427 was linked to the expression of *ABT1* (activator of basal transcription 1), which demonstrated a significant association with hippocampal asymmetry in women. *ABT1* plays a role in transcriptional regulation and may influence hippocampal structure by modulating the gene expression critical for neurodevelopment and neuroplasticity. In addition, *ABT1* has been linked to intelligence,^34^ cognitive ability,^35^ and depression.^36^ Conversely, in men, two distinct CpG sites, cg04564312 and cg06074597, were identified as consistent markers of hippocampal asymmetry. Among these, cg04564312 was associated with the expression of the *RGMB* (repulsive guidance molecule B) gene, which is involved in axonal guidance and neuronal connectivity, processes that may contribute to hippocampal lateralization. These findings suggest that sex-specific epigenetic regulation of key genes, such as *ABT1* and *RGMB*, may underlie differences in hippocampal asymmetry and its evolution over time.

We found evidence for a causal association of cg19045773 with RHCV. This CpG site was linked to the expression levels of the *cis*-genes *ANKRD6* and *RRAGD.* Interestingly, a prior study found a causal relationship between cg26741686 hypermethylation and higher *ANKRD37* gene expression, resulting in the reduction of mean hippocampal volume. ^23^ The Ankyrin repeat domain (ANKRD) family is a widespread protein structural motif with multiple functions, including cell cycle regulation, developmental regulation, cytoskeleton maintenance, and intercellular signaling.^37^ *ANKRD37* is involved in hypoxia, which facilitates the pathogenesis of late-onset Alzheimer’s disease by accelerating Aβ accumulation, increasing tau hyperphosphorylation, impairing the blood-brain barrier, and promoting neuronal degeneration. *ANKRD6* is primarily expressed in neuronal proliferation zones in the brain and has been implicated in brain development.^38^ One recent study found that *ANKRD6* gene expression is linked to sex-specific functional connectivity changes in depression.^39^ *RRAGD* plays a crucial role in the cellular response to amino acid availability through regulation of the mTOR signaling pathway and has been linked to kidney tubulopathy and cardiomyopathy.^40^ Another suggested causal CpG site, cg02929052, whose methylation was associated with RHCV, was associated with lower *UGGT1* and *AMMECR1L* gene expression. UGGT1 is an enzyme involved in the quality control of glycoprotein folding within the endoplasmic reticulum. Dysregulation of these proteins may contribute to the accumulation of toxic protein aggregates. Studies have found that *UGGT1* and other N-glycan modification enzymes are colocalized with Aβ plaques and neurofibrillary tangles in AD brains, suggesting a role in driving glycoprotein remodeling and AD pathogenesis.^41^

Our integrative analysis of methylation, transcription factors and gene expression revealed several TFs that interact with LHCV/RHCV-associated CpGs to co-regulate target gene expression. These findings provide insights into how interactions between the methylome and other molecules contribute to observed phenotypes. Importantly, TF-target associations for these methylation-sensitive TFs often appear only in a subset of samples with high (or low) methylation levels, potentially being overlooked in analyses that aggregate all samples. Recent studies have consistently demonstrated that CpG methylation has a major effect on TF-DNA binding in gene expression regulation.^21,22^ While many of these TFs have been previously linked to neurodegeneration or aging, our integrative analysis offers novel insights into their specific roles in transcriptional regulation. Additionally, we identified target genes for these TFs in the hippocampus, highlighting potential TF-target gene interactions mediated by DNA methylation. For instance, we identified that methylation at cg25770783 and the transcription factor FOXO4 jointly regulate *DHRS1* gene expression related to LHCV. *DHRS1* encodes a member of the short-chain dehydrogenases/reductases (SDR) family, which catalyzes the reduction of steroids as well as prostaglandin E1, satin and xenobiotics, participating in steroid and/or xenobiotic metabolism.^42^ FOXO4 is one of the fundamental anti-stress signaling molecules involved in metabolic regulation, cell survival and proliferation differentiation, DNA damage repair and resilience. In humans, FOXO family and their downstream effectors are thought to be critical in reducing inflammation and is a potential nutraceutical approach to healthy aging and lifespan extension.^43^ In addition, we found that two DMRs and TF ZNF354C jointly regulate *GPR52 and OTUD7A* gene expression, which was associated with hippocampal volume. Previous studies have found that ZNF354C was highly expressed in the brain, including prefrontal cortex, hippocampus and amygdala, and its high expression in hippocampus has been linked to the onset of depression.^44^ *GPR52* plays important roles in signal transduction from the external environment to the inside of the cell, and may impact locomotor activity trough modulation of dopamine, NMDA and ADORA2A-induced locomotor activity.^45^ OTUD7A protein acts on TNF receptor associated factor 6 to control nuclear factor kappa B expression and is an is an emerging independent psychiatric and neurodevelopmental disorders risk gene.^46,47^

While this study provides novel insights into the molecular pathways underlying hippocampal volume and asymmetry, several limitations should be considered. First, our omics data were derived from blood samples, which may not comprehensively capture brain-specific methylation patterns. Although brain tissue is ideal for studying brain-related phenotypes, it is not currently feasible to obtain large-scale methylation data from brain tissues from living human subjects. To address this, we extrapolated our findings by assessing the expression of the identified genes across brain tissues, however, direct comparisons between blood and brain DNA methylation patterns remain limited. Second, while we employed MR analyses to investigate causal relationships, MR has its limitations. For example, it assumes that genetic instruments are independent of confounders and that there is no horizontal pleiotropy,^48^ which can be difficult to fully ensure when assessing in complex traits like brain volumetric measures. Third, each cohort used different brain imaging segmentation algorithm, which could introduce bias. Nevertheless, the quantified brain volumetric measures remained comparable across cohorts, supporting the robustness of our findings. Fourth, our study population was predominantly from European descent, which may limit the generalizability of our findings to other ethnic groups.

In conclusion, our findings provide compelling evidence that DNA methylation signautures in peripheral blood are associated with hippocampal volume, asymmetry and rate of atrophy, underscoring the critical role of epigenetic regulation in brain structure. The methylation signatures identified in this study may serve as potential blood-based biomarkers or therapeutic targets for age- or neurodegeneration-related hippocampal atrophy.

## Methods

### Study participants and analytical overview

The sample included 8,156 participants of European ancestry (EA) from six population-based cohort studies: the Rhineland Study, the Study of Health in Pomerania (SHIP-Trend), the Framingham Heart Study (FHS) offspring study, the Lothian Birth Cohort (LBC) 1936, the Leiden Longevity Study (LLS), and the Older Australian Twin Study (OATS). Participants with known dementia, Parkinson’s disease, prevalent stroke, intracranial tumors, a history of severe head injury, seizures beginning before the age of 25, epilepsy, or multiple sclerosis at the time of MRI scanning were excluded. Details about the included studies and study-specific ethics statements are provided in **Table S1**. Each study obtained written informed consent from all participants and was approved by the appropriate institutional review boards.

### Brain image acquisition and segmentation

Brain MRI acquisition was taken in the same or the closest subsequent visit to the visit in which DNA methylation sample was taken. In each study, MRI scans were acquired and post-processed using standardized procedures without reference to demographic or clinical information. The field strength of the scanners used ranged from 1.5 to 3.0 Tesla. T1- and/or T2-weighted, and/or FLAIR scans were obtained for all participants. All studies used fully automated segmentation methods to quantify brain imaging phenotypes (i.e. LHCV, RHCV, LGMV, RGMV and estimated total intracranial volume (eTIV)). Hippocampal asymmetry and global gray matter asymmetry were defined as the differences between left and right volumes dived by their sums. MRI procedures and quantification in each study are detailed in **Table S2**.

### DNA methylation profiling

Genomic DNA was extracted from blood in each cohort according to standard protocols. Levels of DNA methylation were quantified using the Illumina Infinium MethylationEPIC or Methylation450K BeadChip array. Each cohort performed the quality control for DNA methylation data independently, complying with the agreed quality control guidelines (**Table S3**). The methylation level at each site was represented and analyzed as a β-value, defined as the intensity of the methylated signal/(intensity of the unmethylated signal + intensity of the methylated signal + 100). A β-value of 0 represents a completely unmethylated CpG site and a β-value approaching 1 represents a fully methylated CpG site.

### Statistical analysis

Our workflow to identify novel methylation signatures of LHCV, RHCV, and hippocampal asymmetry is presented in **Fig. 1**. The integrative multi-omics analyses (step 3) and follow-up analyses (step 4) were performed using individual-level data from the Rhineland Study. The overview of the datasets used in the analyses is presented in **Fig. S1**.

#### Cohort-level epigenome-wide association analyses (EWAS)

We quantified the association between DNA methylation level (β-value, independent variable) and each brain imaging-derived endophenotype (outcome) using multivariable linear regression models. Model 1 was adjusted for age, sex, estimated or measured blood cell type proportions (%), ancestry-specific genetic principal components (PCs) and technical covariates (i.e. chip, chip position, and control probe PCs). When hippocampal volumes were outcomes, we additionally adjusted for eTIV. Model 2 was additionally adjusted for smoking status and education. We further adjusted for handedness as a sensitivity analysis (Model 3). To explore whether sex modifies the relationship between DNA methylation and imaging-derived endophenotypes, we also performed sex-stratified analyses. Covariates assessment in each cohort is detailed in **Table S3**.

#### Epigenome-wide meta-analysis

We combined EWAS results based on the sample size-weighted random-effect method with the METAL software.^49^ We additionally performed sex-stratified meta-analyses. Study-specific results were corrected for genomic inflation during meta-analysis if the genomic inflation factor (λ) was larger than one. An association was considered as epigenome-wide significant ^50,51^ if the p-value < 1.0 x10^-7^. CpGs on sex chromosomes and cross-reactive CpGs were removed from the results *post hoc*.

#### Differentially methylated regions analysis

DNA methylation clusters at regions formed by spatially correlated CpGs, namely differentially methylated regions (DMRs), often occur within regulatory regions in the genome and are powerful means to control gene expression.^52^ To account for this, we performed a DMR analysis to identify a group of methylation sites that collectively influence imaging measures using the Comb-p method.^53^ Briefly, Comb-p detects regional enrichment of low p-values at varying distance using the Stouffer-Liptak-Kechris correction for adjacent p-values. A DMR was considered significant if the Šidák corrected p-value < 0.05.

#### Gene set enrichment analysis of identified CpGs/DMRs

To explore the biological pathways underlying the effects of methylation on brain imaging measures, the identified methylation signatures were examined for enrichment in gene sets from the gene ontology (GO) and KEGG databases using missMethyl R package.^54^ The analyses were performed for each imaging measure separately.

#### Look-up analysis of identified CpGs in previous EWASs and GWASs

To identify whether the differential CpGs were associated with other traits, we looked up CpGs showing associations with each imaging measure at epigenome-wide significance level using the EWAS Catalog (http://ewascatalog.org/). We also performed a look-up of known associations of the mapped gene for each CpG and DMR in previously published GWAS using the GWAS catalog (https://www.ebi.ac.uk/gwas).

#### Effects of identified CpGs/DMRs on gene expression levels

Effects of identified CpGs on gene expression for mapped genes and *cis*-genes (± 5Mb)^55^ were investigated in 2,624 participants of the Rhineland Study for whom DNA methylation and gene expression data were available. Detailed gene expression quantification and the QC procedure is described in the **Supplementary methods**. We investigated the association of identified CpGs with their mapped genes and *cis*-gene expression levels using linear regression models adjusted for age, sex, the first 10 genetic PCs, and methylation and gene expression batch effects. We set the statistical significance threshold at p-value < 0.05. The analysis of DMRs was performed similarly, except by replacing CpG methylation levels with the median methylation level of all CpGs located within the DMR.^56^ As the DMR analysis already took the nearby regions into account, we restricted the gene expression analysis to the DMR-mapped genes.

#### Association of gene expression with corresponding imaging measures and mediation analysis

For the genes whose expression levels were associated with CpGs/DMRs, we also quantified the association between gene expression levels and the corresponding imaging measures. Additionally, we evaluated to what extent gene expression levels mediated the effects of CpGs/DMRs on the respective imaging measures.

#### Tissue specificity

To investigate whether our findings were relevant for the brain, we examined the tissue specificity of genes significantly associated with both CpGs or DMRs and the corresponding imaging measures using the Functional Mapping and Annotation (FUMA) of Genome-Wide Association Studies tool.^57^

#### Integrative analysis of identified CpGs/DMRs with putative transcription factors and target genes

Transcription factors (TFs) are proteins that bind to DNA to facilitate transcription, and their binding to DNA can be affected by DNA methylation levels.^21^ To better understand the regulatory roles of the identified CpGs/DMRs on gene expression, we next performed an integrative analysis of DNA methylation, TFs, and gene expression data. We prioritized CpG/DMR-TF-target gene triplets in which regulatory activities of the TFs on target gene expression are most likely influenced by methylation using the *MethReg* R package (v.1.14.0).^58^ Analysis details are described in the **Supplementary methods**.

#### Genome-wide association analysis of identified CpGs and bidirectional two sample Mendelian Randomization analysis

To determine the relationship between genetic variation and methylation levels, known as methylation quantitative trait loci (meQTLs), we performed GWASs of the identified CpGs in 6,723 participants of the Rhineland Study in whom both genetic and methylation data were available. Detailed genetics quantification and the QC procedure are described in **Supplementary methods**. The GWAS of each identified CpG was adjusted for age, sex, methylation data batch effects, smoking status and the first ten genetic PCs to account for population structure. The genome-wide significance level was set at p-value < 5e-8.

Next, we performed bidirectional two-sample Mendelian Randomization (MR) analyses to explore potential causal relationships between the identified CpGs and LHCV/RHCV. In the forward MR analysis, genetic proxies for the identified CpGs were used as the exposure, while genetic proxies for LHCV/RHCV served as the outcome. In the reverse MR analysis, genetic proxies for LHCV/RHCV were used as the exposure, with genetic proxies for the identified CpGs as the outcome. For the CpGs, our GWAS summary statistics were used. For LHCV and RHCV, the UK

Biobank GWAS summary statistics (n=39,691 samples) were used.^59^ The inverse variance-weighted (IVW) method was used as the primary approach for causal inference. Other MR methods including the weighted median and MR Egger method were applied to assess the robustness of the IVW-based MR estimates. The presence of pleiotropy was assessed through the MR Egger intercept test (p-value <0.05). Analysis details are described in the **Supplementary methods**.

#### Association between diet quality scores and identified CpGs/DMRs

As lifestyle factors are one of the main determinants of methylation changes, we also assessed the association between ten diet quality scores and the identified methylation signatures in 5,768 participants of the Rhineland Study in whom both dietary and methylation data were available. We included the following diet quality scores in the analysis: Mediterranean-style diet score (MDS), Dietary Approaches to Stop Hypertension (DASH), Mediterranean–DASH Intervention for Neurodegenerative Delay (MIND) diet, the Alternate Healthy Eating Index (AHEI), the Nordic diet score, EAT-Lancet, plant-based diets as assessed by Plant-based Diet Indexes (i.e. overall PDI, healthful PDI, and unhealthful PDI) and Dietary Inflammatory Index (DII). We used a semi-quantitative Food Frequency Questionnaire (FFQ) to assess participants’ dietary intake, which served as the basis for estimating adherence to the diet quality scores. Further details are provided in the **Supplementary Methods**.

#### Association of identified baseline CpGs/DMRs with longitudinal change in imaging measures

To investigate whether the identified baseline methylation signatures were associated with longitudinal changes in brain imaging measures, linear mixed-effect models were applied to data from 2892 Rhineland Study participants who had complete baseline methylation and follow-up imaing data. Analysis details are provided in the **Supplementary methods**.

## Data availability

The data supporting the findings of this study are included in this manuscript. The complete EWAS summary statistics will be publicly available upon publication.

## Acknowledgement

The authors thank the staff and participants of Rhineland Study, SHIP-Trend, FHS, LBC1936, LLS, and OATS cohorts for their pivotal contributions. We acknowledge collaborative contributions from the NeuroCHARGE working group. A full set of study-specific acknowledgement and funding sources are provided in the **Table S1**.

## Supplementary Materials

Supplementary Figures 1-12

Supplementary Tables 1-8

Supplementary Methods

Supplementary References

