## Supplementary Figures and Methods for "Integrative cross-omics analysis identifies DNA methylation signatures associated with bilateral hippocampal volume, asymmetry and atrophy rate in the general population"

**D. Liu et al.**

**Supplementary Materials**

1. **Supplementary Figures**

**Figure S1:** Overview of the Rhineland Study datasets used in the integrative multiomics and follow-up analyses
**Figure S2:** Distribution of hippocampal and global gray matter volume and asymmetry across age and sex in the Rhineland Study
**Figure S3:** Manhattan plots of the epigenome-wide meta-analyses of hippocampal and global gray matter volume and asymmetry adjusted for handedness
**Figure S4:** Identified CpGs and mapped genes overlap among traits within the model
**Figure S5:** The overlap between genes detected by differentially methylated region analysis among hippocampal and gray matter volume and asymmetry traits
**Figure S6:** Shared gene ontology pathways identified for hippocampal and gray matter volume and asymmetry
**Figure S7:** Specific gene ontology pathways identified for hippocampal and gray matter volume and asymmetry
**Figure S8:** KEGG pathways identified for hippocampal and gray matter volume and asymmetry
**Figure S9:** Genomic annotation of identified methylation signatures
**Figure S10:** The association of CpGs/DMR with putative transcription factors and target gene expressions Manhattan plot showing the genome-wide signals of hippocampal-related CpGs
**Figure S11:** Manhattan plot showing the genome-wide signals of hippocampal-related CpGs
**Figure S12:** Bidirectional two sample Mendelian Randomization analyses reveal causal relationships between identified CpG and right hippocampal volume

2. **Supplementary methods**
3. **Supplementary references**

4. **Supplementary tables:**
   Table S1-S8: separate excel file

**Figure S1. Overview of the Rhineland Study datasets used in the integrative multiomics analysis and follow-up analyses**

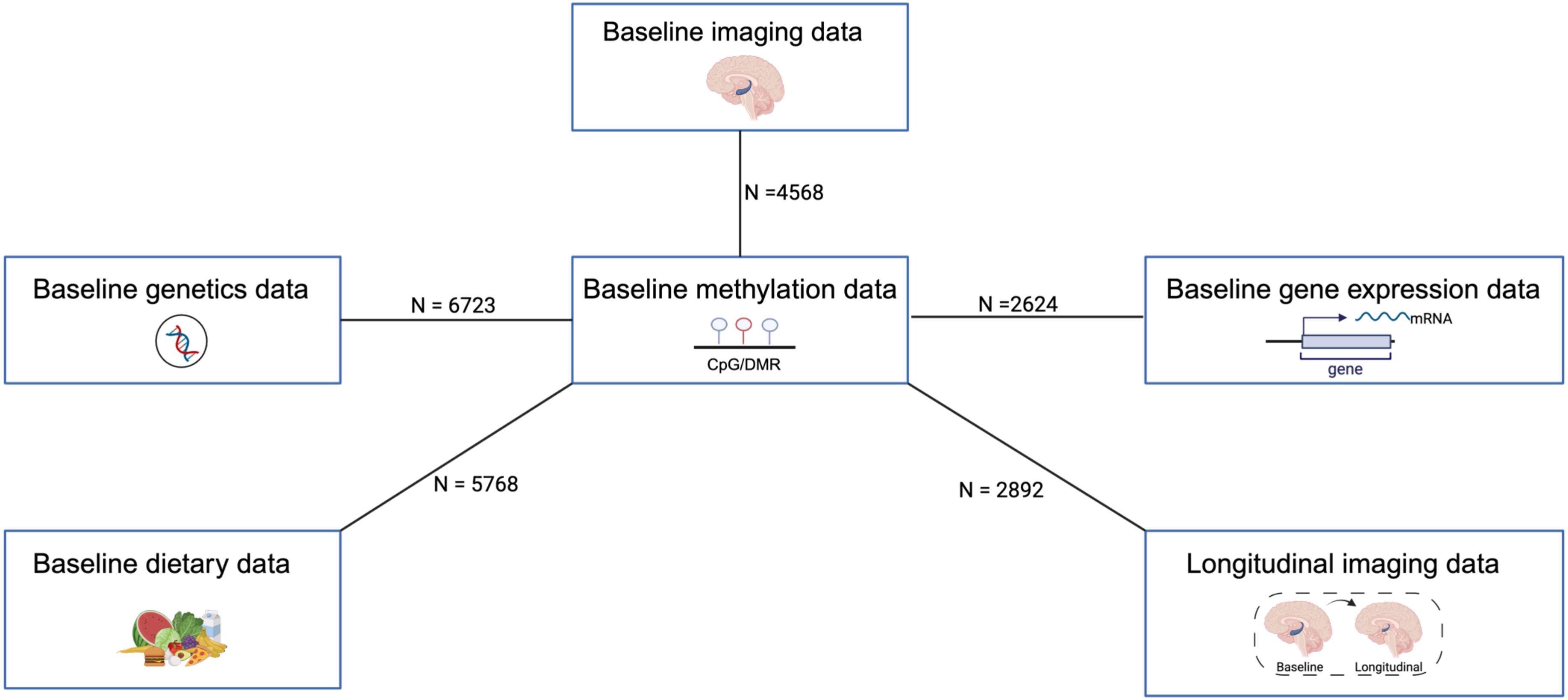


The numbers indicate the sample size available for the assessement of each assocation.

#### Figure S2. Distribution of hippocampal and global gray matter volume and asymmetry across age and sex in the Rhineland Study

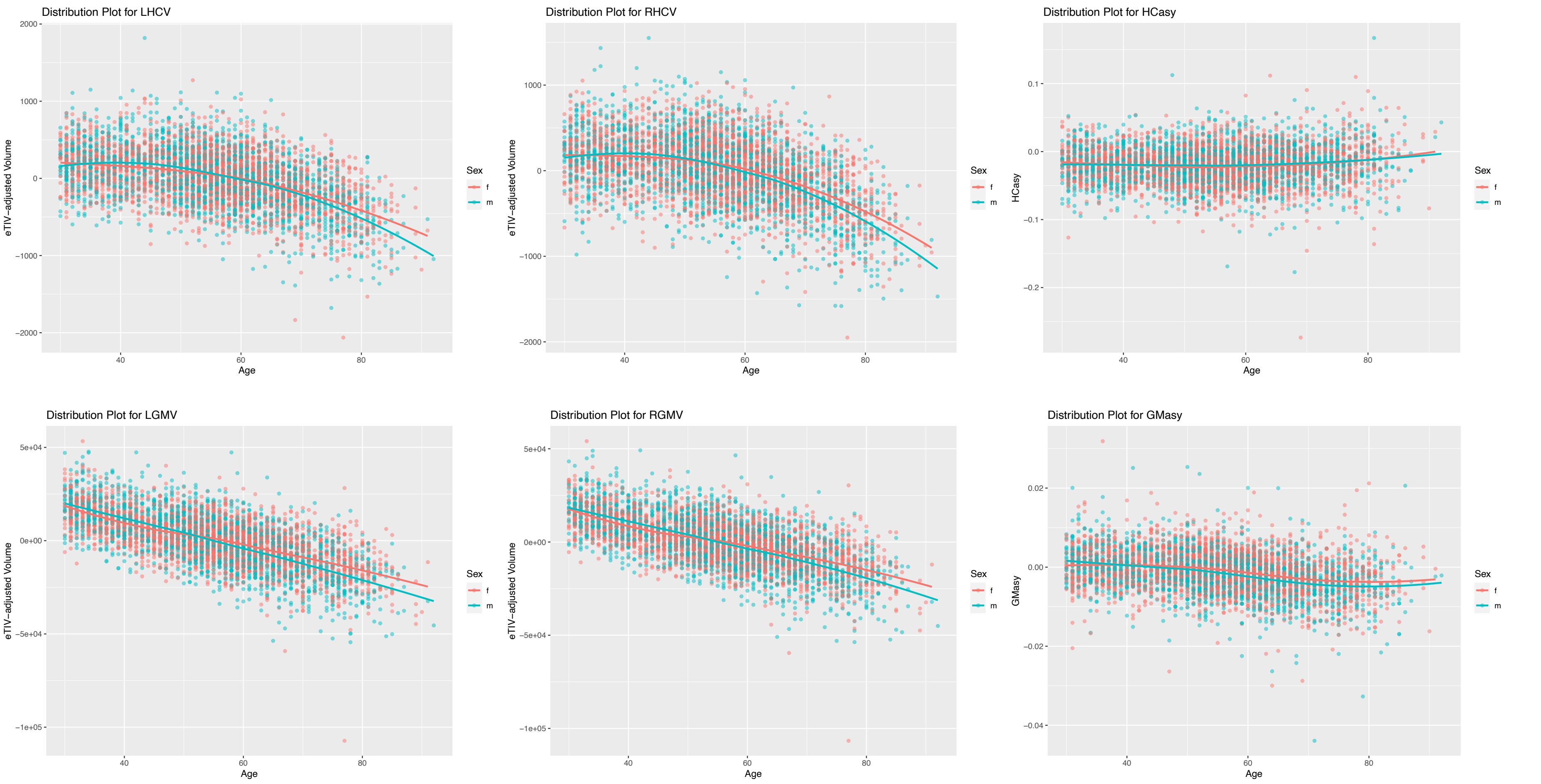


Scatterplots of associations between brain-imaging derived endophenotypes and age. Regression lines for volumetric measures were adjusted for age, sex and estimated total intracranial volume. Regression lines for asymmetry traits were adjusted for age and sex.

Abbreviations: f, female; m, male; LHCV and RHCV, left and right hippocampal volumes; LGMV and RGMV, left and right hemisphere gray matter volumes; HCasy, hippocampal asymmetry; GMasy, global gray matter asymmetry; eTIV, estimated total intracranial volume.

### Figure S3. Manhattan plots of the epigenome-wide meta-analyses of hippocampal and global gray matter volume and asymmetry adjusted for handedness

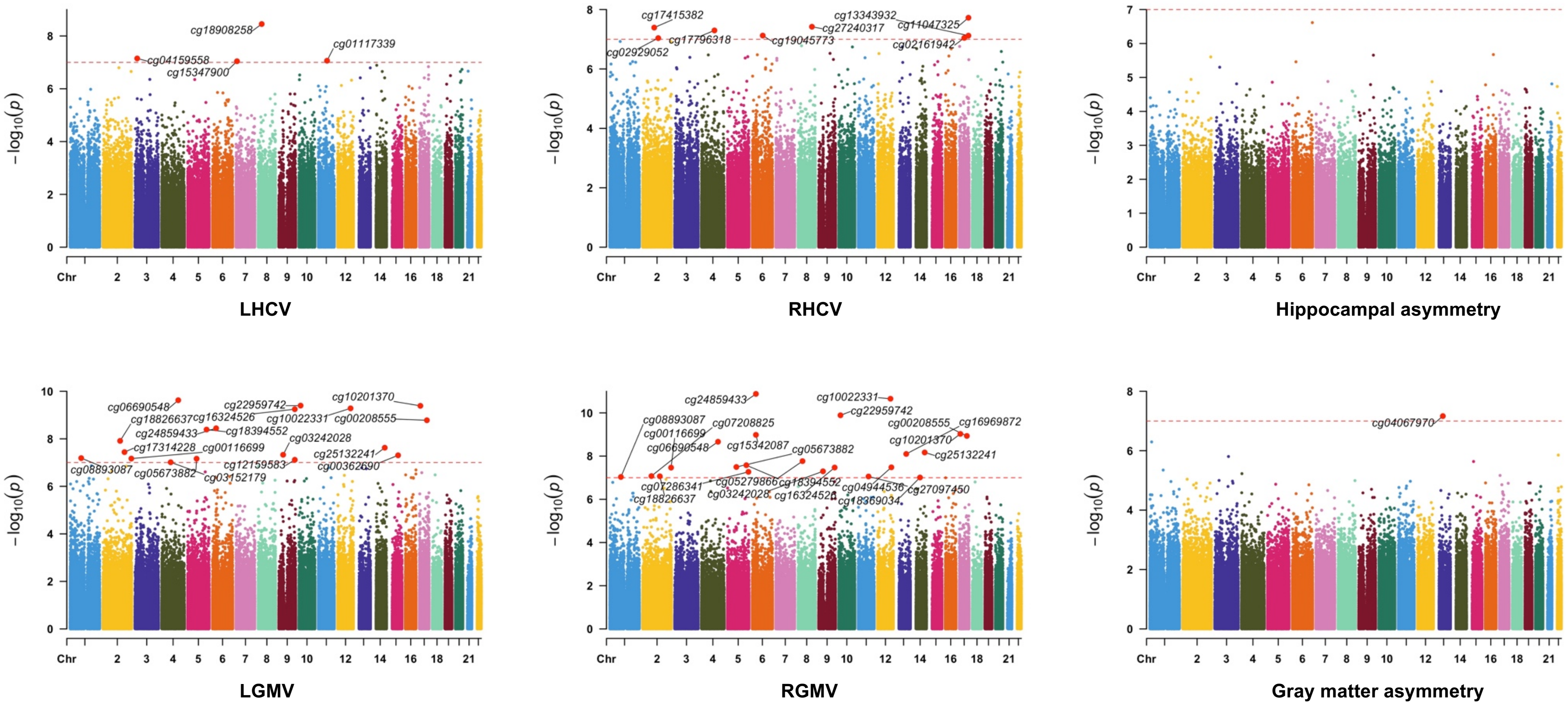


Results were plotted as negative log-transformed p-values (y-axis) across the genome (x-axis). The red horizontal line represents the epigenome-wide significance at 1.0 x10$^{-7}$. Linear models were adjusted for age, sex, estimated total intracranial volume, batch effects, blood cell proportion, the first ten genetic principal components (to account for population stratification), smoking status, education and handedness. All the epigenome-wide significant CpGs were annotated.

Abbreviations: LHCV and RHCV, left and right hippocampal volumes; LGMV and RGMV, left and right hemisphere gray matter volumes.

**Figure S4. Identified CpGs and mapped genes overlap among traits within the model**

A. Identified CpGs/mapped genes overlap among traits

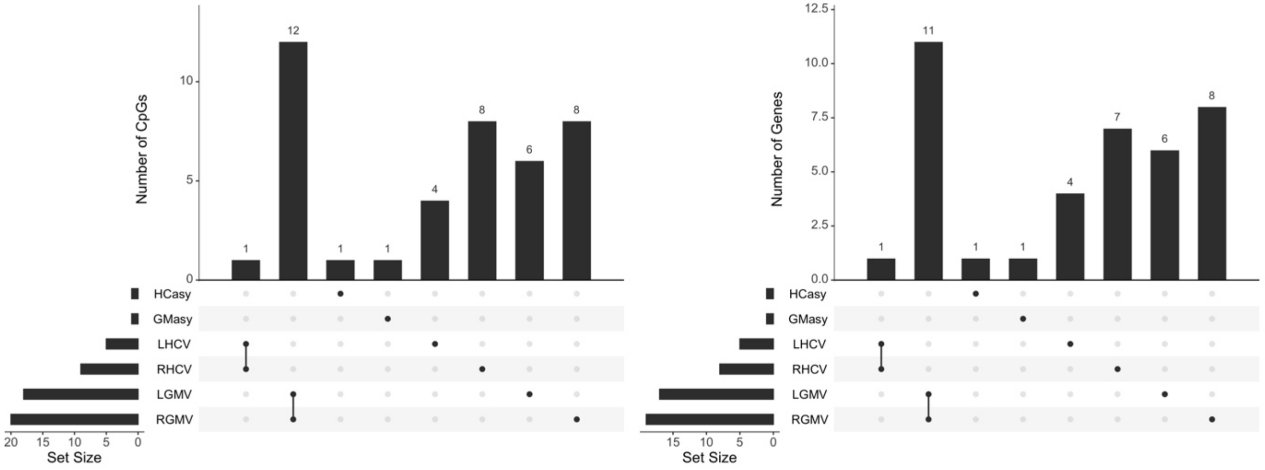


B. Identified CpGs/mapped genes overlap among traits adjusted for handedness

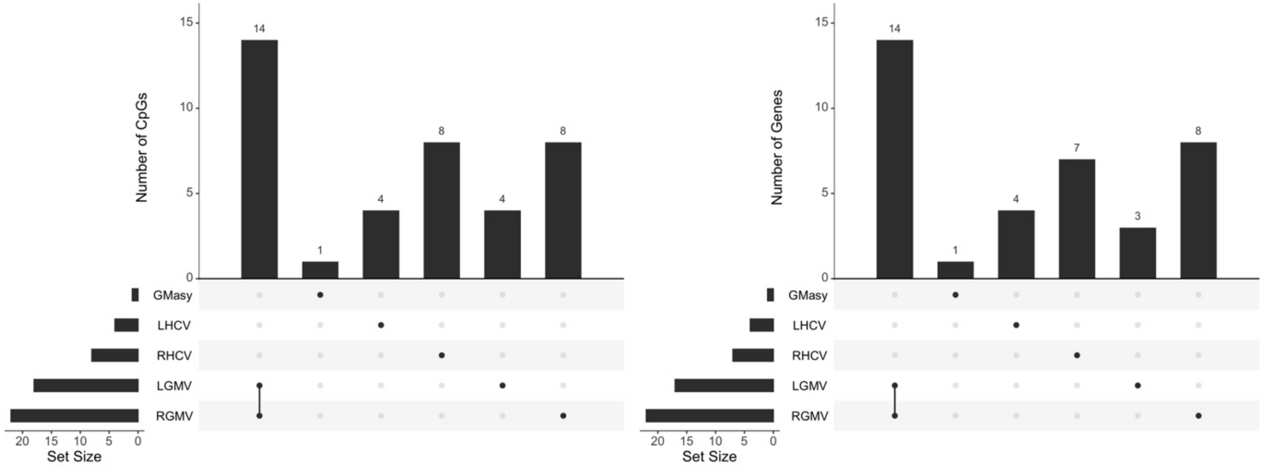


Abbreviations: LHCV and RHCV, left and right hippocampal volumes; LGMV and RGMV, left and right hemisphere gray matter volumes; HCasy, hippocampal asymmetry; GMasy, global gray matter asymmetry.

**Figure S5. The overlap genes between genes detected by differentially methylated region analysis among hippocampal and gray matter volume and asymmetry**

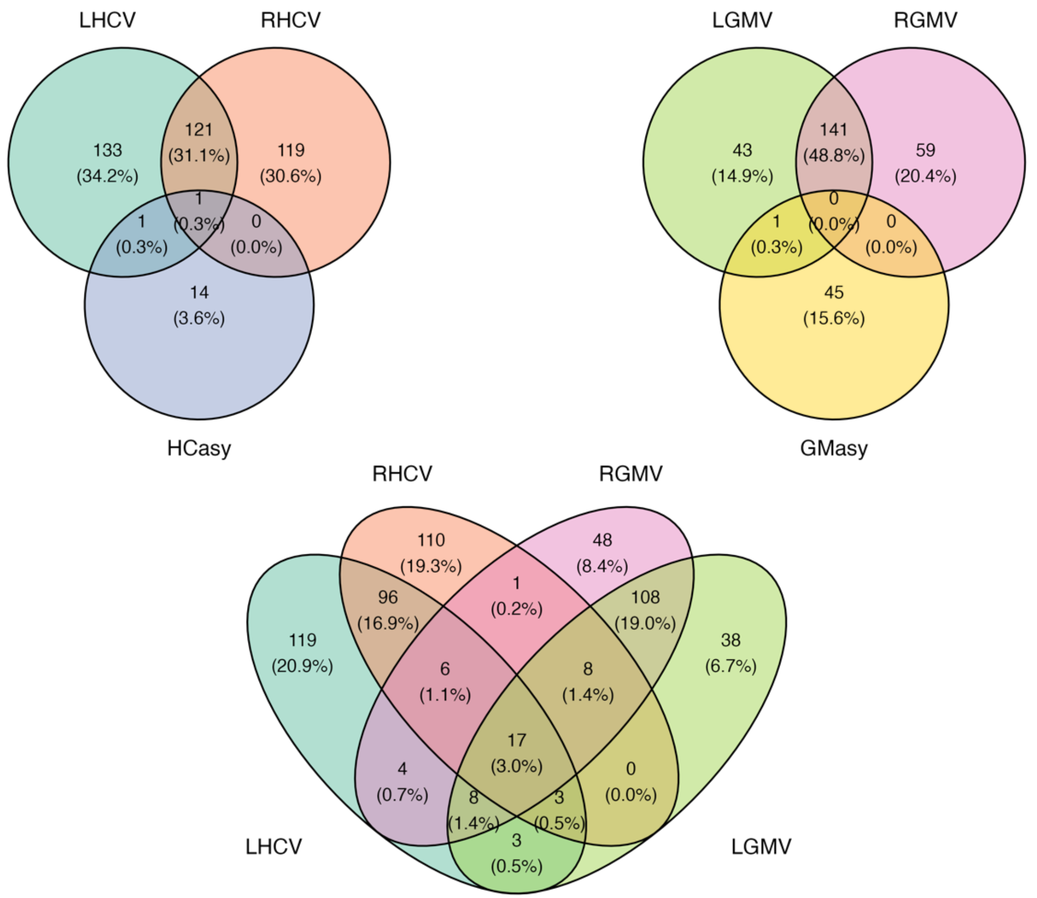


Abbreviations: LHCV and RHCV, left and right hippocampal volumes; LGMV and RGMV, left and right hemisphere gray matter volumes; HCasy, hippocampal asymmetry; GMasy, global gray matter asymmetry.

#### Figure S6. Shared gene ontology pathways identified for hippocampal and gray matter volume and asymmetry

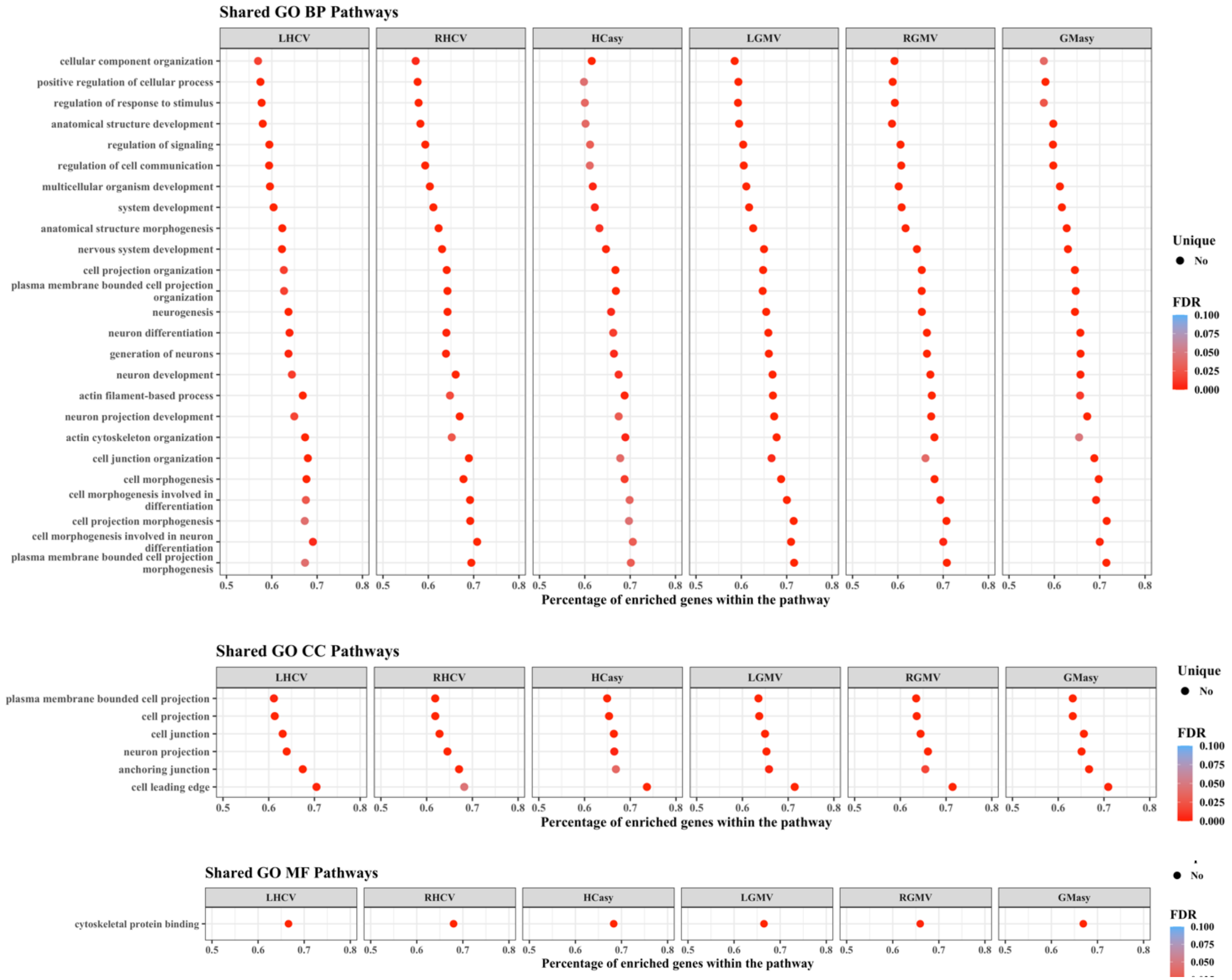


Abbreviations: LHCV and RHCV, left and right hippocampal volumes; LGMV and RGMV, left and right hemisphere gray matter volumes; HCasy, hippocampal asymmetry; GMasy, global gray matter asymmetry. GO: gene ontology database

#### Figure S7. Specific gene ontology pathways identified for hippocampal and gray matter volume and asymmetry

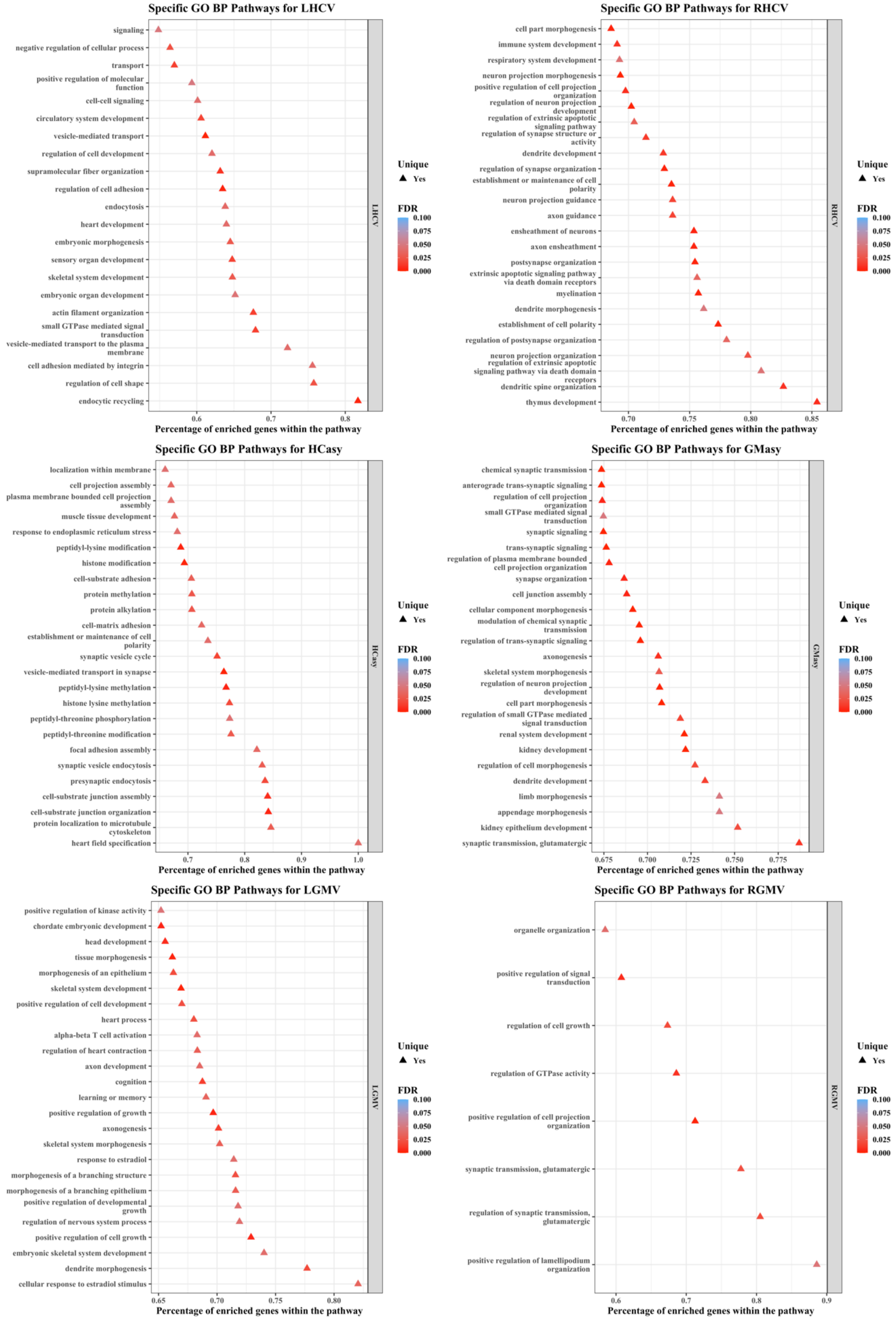


Abbreviations: LHCV and RHCV, left and right hippocampal volumes; LGMV and RGMV, left and right hemisphere gray matter volumes; HCasy, hippocampal asymmetry; GMasy, global gray matter asymmetry; GO BP: gene ontology biological process database; FDR, false discovery rate.

### Figure S8. KEGG pathways identified for hippocampal and gray matter volume and asymmetry

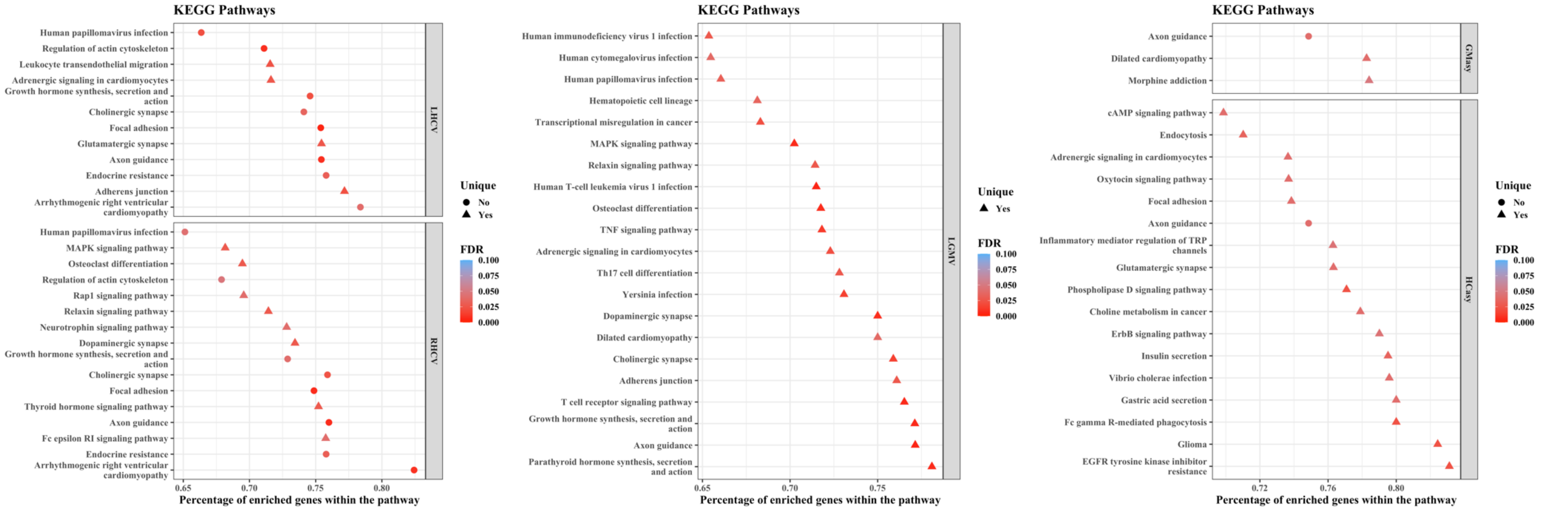


Abbreviations: LHCV and RHCV, left and right hippocampal volumes; LGMV and RGMV, left and right hemisphere gray matter volumes; HCasy, hippocampal asymmetry; GMasy, global gray matter asymmetry; FDR, false discovery rate; KEGG, Kyoto Encyclopedia of Genes and Genomes;

#### Figure S9. Genomic annotation of identified methylation signatures

A. LHCV

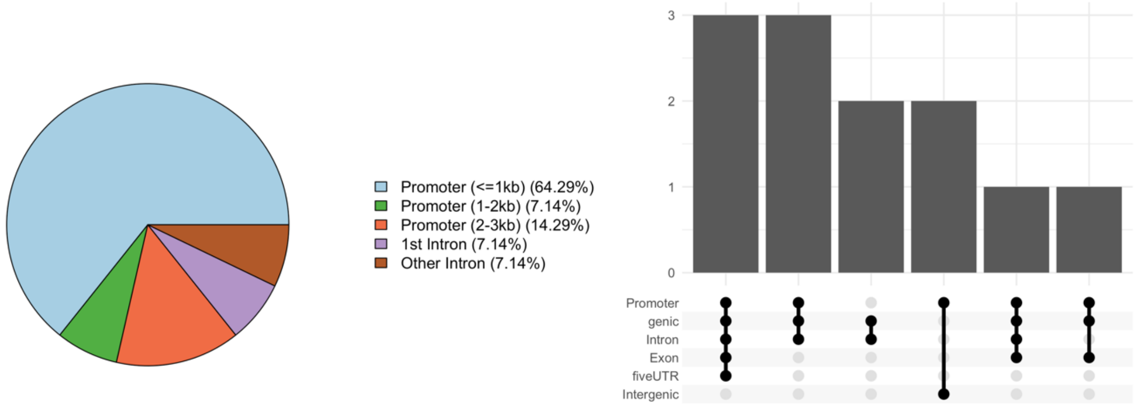


B. RHCV

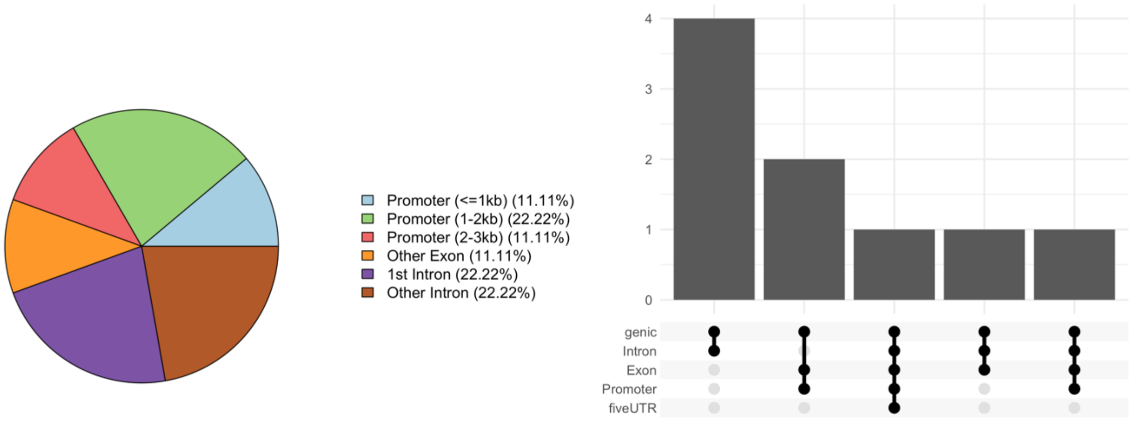


Genomic annotation of CpGs and DMRs loci associated with LHCV (A) and RHCV (B). The pieplots and the upset plots were generated using ChIPseeker v1.26.2 R package.
Abbreviations: LHCV and RHCV, left and right hippocampal volumes.

### Figure S10. The association of CpGs/DMR with putative transcription factors and target gene expressions

**A**

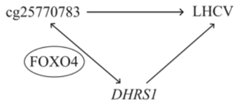


| | |
|---|---|
| Genome of reference | hg19 |
| Region ID | chr14:24551142-24551143 |
| Probe ID | cg25770783 |
| Target gene ID | ENSG00000157379 |
| Target gene Symbol | DHRS1 |
| TF gene ID | ENSG00000184481 |
| TF gene Symbol | FOXO4 |
| TF role | Activator |
| DNAm effect | Attenuating |

| Target ~ TF + DNAm Quant. Group + TF * DNAm Quant. Group | Estimate | P-Values |
|---|---|---|
| Direct effect of DNAm | -0.0194 | 0.672226 |
| Direct effect of TF | 0.165 | 1.77e-06 |
| Synergistic effect of DNAm and TF | -0.177 | 0.000247 |

| Legend |
|---|
| rlm: robust linear model |

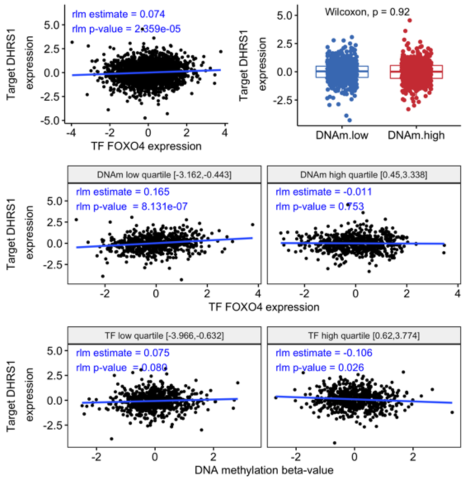


**B**

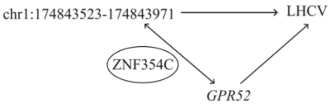


| | |
|---|---|
| Genome of reference | hg19 |
| Region ID | chr1:174843523-174843971 |
| Target gene ID | ENSG00000203737 |
| Target gene Symbol | GPR52 |
| TF gene ID | ENSG00000177932 |
| TF gene Symbol | ZNF354C |
| TF role | Repressor |
| DNAm effect | Enhancing |

| Target ~ TF + DNAm Quant. Group + TF * DNAm Quant. Group | Estimate | P-Values |
|---|---|---|
| Direct effect of DNAm | 0.191 | 1.77e-05 |
| Direct effect of TF | -0.552 | < 2e-16 |
| Synergistic effect of DNAm and TF | 0.143 | 0.005752 |

| Legend |
|---|
| rlm: robust linear model |

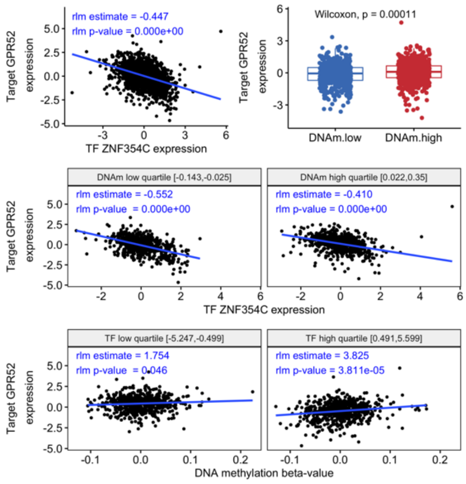


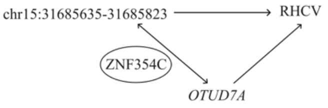


| | |
|---|---|
| Genome of reference | hg19 |
| Region ID | chr15:31685635-31685823 |
| Target gene ID | ENSG00000169918 |
| Target gene Symbol | OTUD7A |
| TF gene ID | ENSG00000177932 |
| TF gene Symbol | ZNF354C |
| TF role | Repressor |
| DNAm effect | Attenuating |

| Target ~ TF + DNAm Quant. Group + TF * DNAm Quant. Group | Estimate | P-Values |
|---|---|---|
| Direct effect of DNAm | -0.01 | 0.819111 |
| Direct effect of TF | -0.185 | 4.98e-07 |
| Synergistic effect of DNAm and TF | 0.203 | 7.82e-05 |

| Legend |
|---|
| rlm: robust linear model |

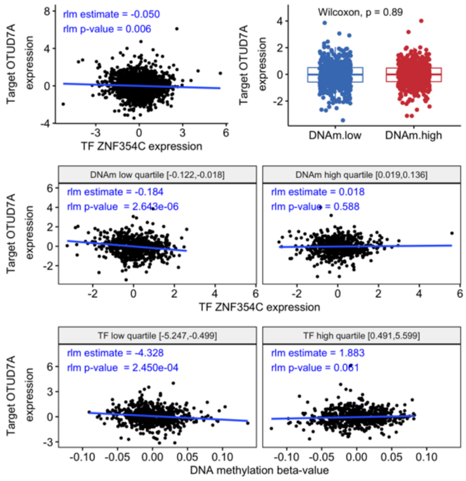


Results of MethReg analysis. The plots show the relationship between DNA methylation and target gene (1st row), the relationship between transcription factor and target genes in low and high DNA methylation level groups (2nd row) and the relationship between DNA methylation and target gene in low and high transcription factor level groups (3rd row). (A) Results for cg25770783 – FOXO4 – DHRS1 triplet for which the interaction term between cg25770783 and FOXO4 was significant. cg25770783 attenuated FOXO4 activity. (B) Results for two triplets for which the interaction terms between DMR and transcription factor, as well as the association between the target gene and the related trait, were significant. In the “chr1:174843523-174843971 - ZNF354C - GPR52” triplet, the DMR enhances the transcription factor activity. In the “chr15:31685635-31685823 - ZNF354C - OTUD7A” triplet the DMR attenuates the transcription factor activity.

**Figure S11. Manhattan plot showing the genome-wide signals of hippocampal-related CpGs**

A. GWAS of LHCV-related CpGs

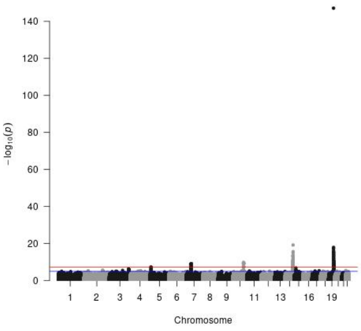


cg03826366

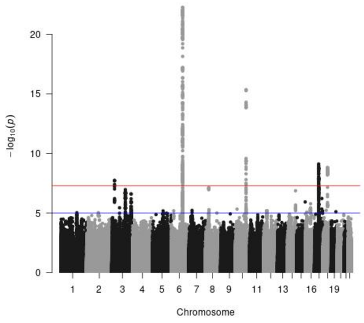


cg13343932

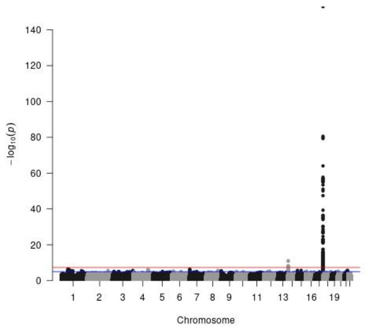


cg25770783

B. GWAS of RHCV-related CpGs

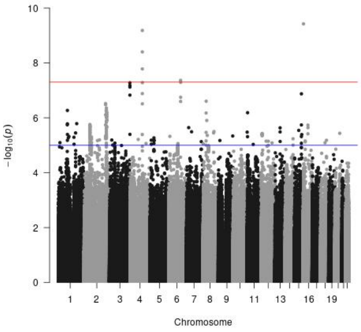


cg02348995

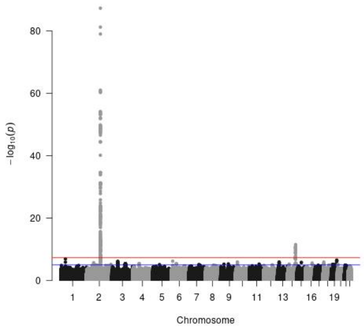


cg02929052

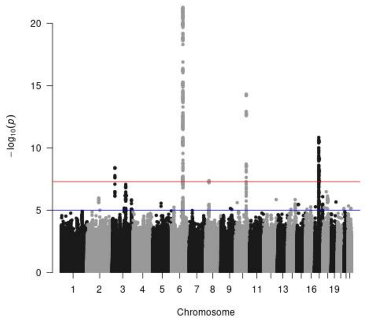


cg11047325

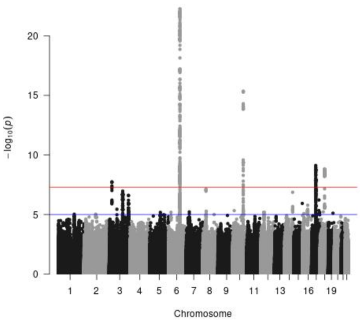


cg13343932

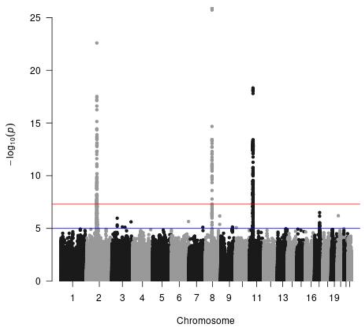


cg17415382

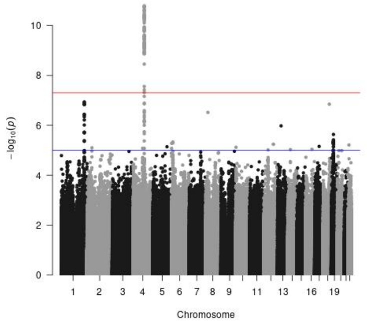


cg17796318

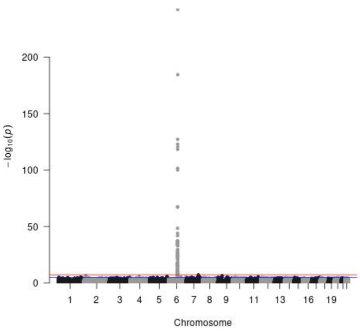


cg19045773

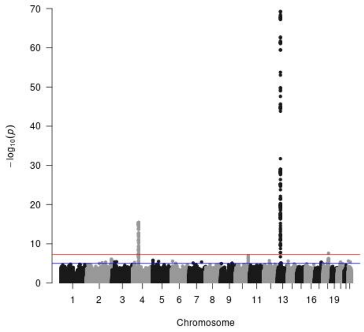


cg19122233

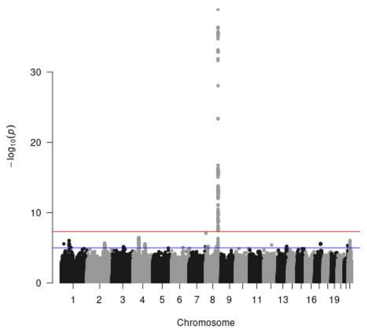


cg27240317

**Figure S12. Bidirectional two sample Mendelian Randomization analyses reveal causal relationships between identified CpG and right hippocampal volume**

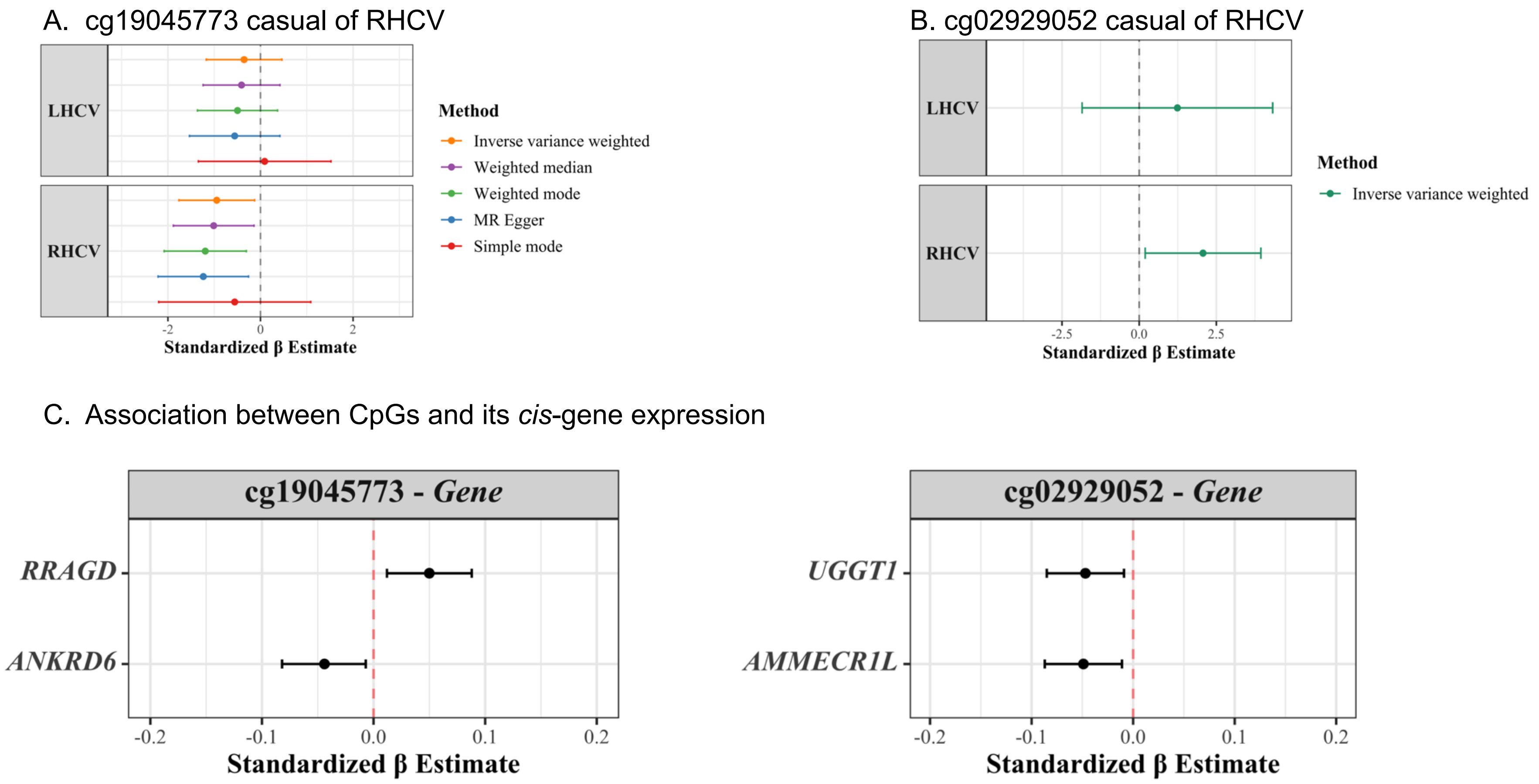


Methylation site cg19045773 and cg0292052 were identified to be specifically associated with RHCV in the EWAS meta-analysis.
(A) Forest plot showing causal effect estimates from the two sample Mendelian Randomization analysis of the effect of cg19045773 on LHCV and RHCV. The dot represents the mean causal effect and the horizontal line shows the 95% CI. (B) Forest plot showing causal effect estimates from the two sample Mendelian Randomization analysis of the effect of cg0292052 on LHCV and RHCV. The dot represents the mean causal effect and the horizontal line shows the 95% CI. (C) Forest plot showing the effect of site cg19045773 and cg0292052 on their cis-gene expressions. The dot represents the mean effect and the horizontal line shows the 95% CI. Abbreviations: LHCV and RHCV, left and right hippocampal volumes.

#### Supplementary Methods

##### Whole blood RNA isolation and gene expression profiling

We extracted total RNA from blood. Blood was collected in PAXgene Blood RNA tubes (PreAnalytix/Qiagen) and processed according to manufacturer's guidelines. PAXgene tubes were thawed and incubated at room temperature to increase RNA yields. Total RNA was isolated according to manufactures's instuctions using PAXgene Blood miRNA Kit and following the automated purification protocol (PreAnalytix/Qiagen). RNA integrity and quantity were evaluated using the Tapestation 4200 instrument (Agilent). We used 750 ng total RNA to generate NGS libraries using the TruSeq stranded total RNA kit (Illumina) following manufacturer's instructions with Ribo-Zero Globin reduction. We quantified the libraries via Qubit HS dsDNA assay (Invitrogen) and clustered at 250 pM final clustering concentration on a NovaSeq6000 instrument using S2 v1 chemistry (Illumina) in XP mode for the first 3,000 samples and NoveSeq S4 v1.5 chemistry for the latest 384 samples, and sequenced paired-end 2*50 cycles before demultiplexing using bcl2fastq2 v2.20. Quality control of the sequencing was evaluated through FastQC (v0.11.9). To quantify the expression of genes in RNA-Seq data, the sequencing reads were first aligned to the human reference genome GRCh38.p13 provided by Ensembl using STAR v2.7.1. The count matrix was generated with STAR –quantMode GeneCounts using the human gene annotation version GRCh38.101. Genes with an overall mean expression greater than 15 reads and expressed in at least 5% of the participants were used for the following analyses. Finally, in order to normalize for library size and to log-transform the raw data, we applied the varianceStabilizingTransformation function from DESeq2 v1.30.1 R package.

##### MethReg integrative analysis

To create CpG-TF-target gene triplets, we first linked a given CpG to transcription factors (TFs) with binding sites within ±250 bp of the CpG, using information from the ReMap2020 database, which contains regulatory regions for over a thousand transcriptional regulators obtained using genome-wide DNA-binding experiments such as ChIP-seq. Next, we linked a CpG to a gene if it was located within the gene's promoter region (< ±2 kb from the transcription start site). For CpGs in distal regions, we linked them either to genes within 500 kb or to the five nearest upstream and five nearest downstream genes from the CpG location. The CpG-TF pairs were then combined with CpG-target gene pairs to create triplets of CpG-TF-target genes. We used the same matched methylation-RNA samples from the Rhineland study. TF activities were estimated using the GSVA R package. The MethReg analyses were performed using the MethReg R package (v.1.14.0).[1]

##### Genomics and bidirectional two-sample Mendelian Randomization (MR)

Blood samples were genotyped using the Illumina Omni-2.5 exome array, which contains 2,612,357 single-nucleotide polymorphisms (SNPs). Subsequently, genotype data were processed using GenomeStudio (v.2.0.5), and quality controlled with PLINK (v.1.9). SNPs were excluded if not meeting the Hardy-Weinberg disequilibrium criterium (p-value < 1E-5), having a minor allele frequency < 0.01, or showing a poor genotyping rate (< 99%). Additionally, participants were excluded because of a poor call rate of less than 95% (n = 51), abnormal heterozygosity (n = 100), cryptic relatedness (n = 472), or a sex mismatch (n = 43). To account for variation in population structure, which may otherwise cause systematic differences in allele frequencies,[2] we used EIGENSTRAT (v.16000). EIGENSTRAT uses principal components to detect and correct for population structure, which resulted in the exclusion of an additional 236 participants from non-Caucasian descent. Finally, we imputed missing SNPs with IMPUTE (v.2)[3] based on the 1000 Genomes (phase 3) reference panel.[4] We ensured a high imputation quality by only including SNPs with an info score metric > 0.3, which indicates reliable imputation.[5]

To determine the relationship between genetic variation and methylation levels, known as methylation quantitative trait loci (meQTLs), we performed GWASs of the identified CpGs in 6,723 participants of the Rhineland Study in whom both genetic and methylation data were available. The GWAS of each identified CpGs was adjusted for age, sex, methylation data batch effects, smoking status and the first ten genetic PCs to account for population structure. The genome-wide significance level was set at p-value < 5e-8.

Next, we performed bidirectional two-sample Mendelian Randomization (MR) analyses to explore potentially causal relationships between the identified CpGs and LHCV/RHCV. The forward MR analyses were performed using the genetic proxies for the identified CpGs as the exposure and those for LHCV/RHCV as the outcome. In the reverse MR analyses, genetic proxies for LHCV/RHCV were used as the exposure and those for the identified CpGs as the outcome. For the CpGs, our GWAS summary statistics were used. For LHCV and RHCV, the UK Biobank GWAS summary statistics (n=39,691 samples) were used.[6] To meet MR assumptions, only SNPs that were strongly associated (p-value $< 5 \times 10^{-8}$) with each exposure were selected. Second, we performed linkage-disequilibrium (LD) pruning to select the independent SNPs. The clump function in the R package *TwoSampleMR* (v.0.5.6) [7] was used for LD pruning with the following parameters: $r^2 = 0.001$, a window size of 1,000 kb, and the European population of the 1000 Genomes Project as the reference panel. The data on exposure and outcome were harmonized with regard to effect directions, and palindromic SNPs with minor allele frequency close to 0.5 were removed.

**Dietary Assessment and Diet Quality Scores**

Habitual dietary intake was assessed by a self-administered semi-quantitative food frequency questionnaire (FFQ). The questionnaire was originally developed for the European Prospective Investigation into Cancer and Nutrition (EPIC) study in Potsdam and adapted for the Rhineland Study. Participants were questioned about their habitual intake of 132 food and beverage items in the last 12 months, including standard portion sizes.[8] Depending on the food item, there were between four and eleven frequency options available, ranging from “never” to “11 times per day or more”. Additionally, participants were also asked about fat contents of consumed dairy and meat products, as well as types of fat used for food preparation.

The FFQs were eligible for inclusion in analyses provided that information was available for at least 80% of core food items. Fats used for food preparation or additives to hot beverages were not considered as core food items. Missing data on the FFQ were found for 118 core food items, with a maximum of 31 missing values in one FFQ item. Most food items had only 1 or 2 missing values. To retain all observations with ≥80% of questionnaire competition, missing values on the FFQ were imputed using MissForest. Using SAS 9.4, we computed the sum intake of each food/beverage and estimated macro- and micronutrients, water, and energy intake based on data from the German Food Code and Nutrient Database[9] (v.3.02). The dietary data were categorized into 17 main food groups and 71 subgroups in accordance with the EPIC SOFT classification scheme.

Diet quality scores including Mediterranean-style diet score (MDS),[10] Dietary Approaches to Stop Hypertension (DASH),[11] Mediterranean–DASH Intervention for Neurodegenerative Delay (MIND) diet,[12] the Alternate Healthy Eating Index (AHEI),[13] the Nordic diet score,[14] EAT-Lancet,[15] plant-based diets as assessed by Plant-based Diet Indexes (i.e. overall PDI, healthful PDI, and unhealthful PDI)[16] and Dietary Inflammatory Index (DII)[17] were caculated following previous published methods.

**Association of identified baseline methylation signatures with longitudinal change in brain imaging measures**

For longitudinal analysis, we used follow-up imaging data. Complete follow-up data was available in 2892 participants. To investigate whether the identified baseline methylation signatures were associated with longitudinal changes in imaging measures, linear mixed-effect models were applied as follows:

$$Y_i = \beta_0 + \beta_1 \cdot age + \beta_2 \cdot CpG/DMR + \beta_3 \cdot age \times CpG/DMR + \; b_{0,i} + b_{1,i} \cdot age + \varepsilon_\iota$$

Here, $Y_i$ denotes the imaging measure at any time point for participant *i*, $\beta_0$ denotes the fixed mean intercept, $\beta_1$ denotes the fixed effect of age (i.e., the average change rate of the outcome per year across all individuals), $\beta_2$ is the fixed effect of baseline methylation signatures, $\beta_3$ is the fixed effect of the interaction between baseline methylation signatures and age, and $\varepsilon_i$ represents the residual error. To account for multiple measurements of the same participant across time, we included a random intercept ($b_{0,i}$) and slope ($b_{1,i}$) for each participant. The effect of baseline methylation signatures on the average change rate of imaging measures was thus given by the interaction term $\beta_3$. The average change rate associated with one standard deviation (SD) increase of baseline methylation signatures was calculated as $\beta_1 + \beta_3$. All available brain imaging examinations from any time point were included in the model and contributed towards the coefficients. As with the cross-sectional analysis, we adjusted the models for age, sex, batches, and in case of hippocampal volume, for eTIV.

To evaluate the variance explained by methylation markers on the random slope, we compared the variance in the random slope of age in a base model, only adjusted for age, sex, and eTIV, to that in the full model, which included all identified CpG or DMR and their interactions with age. All linear mixed effect models were analyzed with the nlme R package. Model variances were extracted using the **VarCorr** function from the same package. The percentage of variance explained was calculated as:

*Explained variance (%) = [1- random slope variance (full model)/random slope variance (base model)]*100*

This approach allowed us to quantify the contribution of baseline methylation signatures to explaining inter-individual differences in the rate of imaging measure changes over time.
